# Perceived Consequences and Catastrophising Help Explain Health-Related Quality of Life in Parkinson’s Disease. A Cross-Sectional Study

**DOI:** 10.64898/2026.04.27.26351802

**Authors:** Viktoria Azoidou, Essa Bhadra, Ellen Camboe, Kamalesh C. Dey, Alexandra Zirra, Kira Rowsell, Corrine Quah, Caroline Budu, Thomas Boyle, David Gallagher, Jonathan P. Bestwick, Laura J Smith, Alastair J Noyce, Cristina Simonet

**Author notes:** Corresponding authors: Viktoria Azoidou, PhD, & Cristina Simonet, PhD, Centre for Preventive Neurology, Wolfson Institute of Population Health, Queen Mary University of London, London, United Kingdom, &.

## Abstract

**Introduction:** Motor complications are major determinants of disability in Parkinson’s disease (PD), yet clinician-rated motor complication severity does not fully explain variability in health-related quality of life (HRQoL).

**Research question:** To examine the contribution of illness perceptions and cognitive-behavioural responses to HRQoL alongside motor complication severity in people with PD.

**Methods:** This multi-centre, cross-sectional study recruited 58 people with idiopathic PD (median age 68 years; 55.2% male; 48.3% from minoritised ethnic backgrounds; Hoehn & Yahr stage 2-3). All underwent assessment of motor complications (Movement Disorder Society-Unified Parkinson’s Disease Rating Scale; MDS-UPDRS Part IV) and HRQoL (Parkinson’s Disease Questionnaire-39 Summary Index; PDQ-39 SI). Illness perceptions were measured with Illness Perception Questionnaire-Revised (IPQ-R) Part 2, and cognitive-behavioural responses with Cognitive and Behavioural Responses Questionnaire (CBRQ). Regression models were adjusted for age, sex, disease duration, motor severity (MDS-UPDRS Part III), levodopa equivalent daily dose (LEDD), anxiety, depression, and cognitive function. A subset (n=47) completed 7-day Parkinson’s KinetiGraph monitoring.

**Results:** Demographic and clinical covariates explained 77.3% of variance in HRQoL (R²=0.773). Adding motor complication severity explained a significant additional 3.7% (ΔR²=0.037, P=0.004). Subsequent inclusion of illness consequences (IPQ-R) and catastrophising (CBRQ) explained a further 4.1% (ΔR²=0.041, P=0.004), yielding a final adjusted R² of 0.815. In the fully adjusted model, catastrophising (B=0.797, P=0.027) and perceived consequences (B=0.767, P=0.013) remained independently associated with HRQoL.

**Conclusion:** HRQoL in PD appears to depend not only on motor complication severity, but also on patients’ interpretations and responses. Clinicians should assess both to guide holistic care and support adaptive coping.

## INTRODUCTION

Health-related quality of life (HRQoL) is a core patient-reported outcome in Parkinson’s disease (PD), reflecting the cumulative impact of motor and non-motor symptoms on function, emotional well-being, autonomy, and social participation[1]. PD is a chronic, progressive neurodegenerative disorder characterised by bradykinesia, rigidity, and tremor, with advancing pathology leading to increasing disability and loss of independence[2]. PD is the second most prevalent neurodegenerative disorder globally and the fastest-growing neurological cause of disability. In 2021, an estimated 11.77 million people were living with PD, representing a 2.7-fold increase since 1990, with projections reaching 25.2 million by 2050[3]. As this burden rises, understanding drivers of HRQoL variability is critical for clinical care and service planning.

Motor complications, particularly motor fluctuations and levodopa-induced dyskinesias (LID), contribute to reduced HRQoL[4]. Approximately 40% of individuals develop motor fluctuations and a similar proportion develop dyskinesias within 4-6 years of levodopa exposure, although estimates vary by cohort and time origin[5]. Longitudinal incident cohorts provide longer-term context: motor fluctuations affect 54.3% of patients by 5 years and nearly all by 10 years, while dyskinesias affect 55.7% by 10 years[6].

The development of motor complications increases treatment complexity and often prompts consideration of device-aided therapies[2,7]. Reducing motor complication burden, particularly following advanced interventions, is associated with clinically meaningful HRQoL improvement[8,9]. However, motor complication severity does not fully explain HRQoL variability[10]. Individuals with comparable clinician-rated impairment frequently report markedly different PDQ-39 outcomes[1,10], even after adjusting for age at onset, sex, disease duration, and dopaminergic exposure[4,11,12].

This discordance suggests a role for psychological factors. Psychological adjustment demonstrates stronger associations than clinical stage with several Parkinson’s Disease Questionnaire-39 (PDQ-39) domains, including emotional well-being and stigma[13]. How individuals respond to stress and illness independently shapes their lived experience of PD[1]. Well-being extends beyond motor control and encompasses autonomy, competence, and participation in meaningful social, occupational, and community roles[14]. Anxiety and depression, affecting 30-50% of people with PD, strongly correlate with poorer HRQoL[10,15,16]. Qualitative studies[17,18] further show that identity, psychosocial, and environmental factors influence HRQoL independently of symptom severity, while perceived independence and continued engagement in valued activities are central to well-being. Unpredictable OFF periods and visible dyskinesias may destabilise identity and autonomy[19], and stigma-related experiences can amplify distress[20].

The Common-Sense Model of self-regulation[21–23] provides a framework linking illness representations to coping and adaptation. Illness perceptions, measured using the Illness Perception Questionnaire-Revised (IPQ-R)[24], assess beliefs about consequences, controllability, chronicity, coherence, and emotional impact. Cognitive-behavioural responses, assessed with the Cognitive and Behavioural Responses Questionnaire (CBRQ)[25,26], capture behavioural patterns such as avoidance, symptom monitoring, and activity regulation. Although related, illness perceptions and behavioural responses operate at distinct levels and may relate differentially to motor complication burden and HRQoL. Given the differing neurobiological and experiential profiles of fluctuations and dyskinesias[2,4,6], their psychological correlates may also diverge.

Sociodemographic context further shapes HRQoL. In US Parkinson’s Foundation Centers of Excellence, minoritised racial and ethnic groups reported poorer PDQ-39 scores than White participants after adjustment for disease severity, with cognitive performance partially mediating differences[27]. Cognitive disparities between Latino and White non-Latino patients have also been documented[28]. In the United Kingdom, the East London Parkinson’s Disease Project found lower EQ-5D-5L utilities among South Asian participants and greater depressive burden and cognitive impairment in South Asian and Black patient groups[29]. These findings emphasise the multidimensional determinants of perceived health status.

Clinician-rated motor complication severity is routinely assessed using Part IV of the Movement Disorder Society-Unified Parkinson’s Disease Rating Scale (MDS-UPDRS)[30], while wearable-derived metrics provide objective measures of free-living motor heterogeneity[31]. However, no study has concurrently examined clinician-rated motor complications, illness perceptions, cognitive-behavioural responses, and wearable-derived motor markers within a unified analytical framework to determine their relative and combined associations with HRQoL.

We conducted a cross-sectional study in a diverse PD cohort to examine how motor complication severity, illness perceptions, and cognitive-behavioural responses relate to HRQoL. We hypothesised that psychological constructs would explain additional variance in HRQoL beyond demographic and clinical variables, including motor severity. Secondary analyses explored associations between motor complications and psychological measures, the interrelationship between illness representations and behavioural responses, and their independent and joint contributions to HRQoL. Wearable-derived metrics were included to determine whether objective free-living motor burden demonstrated similar associations.

## METHODS

### Trial design

This multi-centre, cross-sectional study was embedded within a registered clinical project consisting of a feasibility study (n=10 participants with idiopathic PD) and pilot double-blind randomised controlled trial (n=48 participants with idiopathic PD) both investigating a noninvasive intervention delivering vibrotactile stimulation, undertaken at the Wolfson Institute of Population Health, Queen Mary University of London, in partnership with Barts Health NHS Trust and Homerton Healthcare NHS Foundation Trust. The present analyses draw exclusively on baseline data acquired prior to initiation of any intervention. Ethical approval was granted by the London-Dulwich Research Ethics Committee (23/PR/1526). All procedures conformed to the principles of the Declaration of Helsinki, and written informed consent was obtained from all participants. Study oversight included input from a Patient and Public Involvement and Engagement advisory group. Reporting adheres to STROBE guidance (SUPPLEMENTARY MATERIAL A).

### Participants

Participants were recruited from specialist outpatient clinics at Barts Health NHS Trust (Neurology) and Homerton Healthcare NHS Foundation Trust (Care of the Elderly). Inclusion criteria comprised idiopathic PD diagnosed according to Movement Disorder Society clinical diagnostic criteria[32], age ≥18 years, fluency in English sufficient to complete study assessments, and capacity to provide informed consent. Exclusion criteria included atypical parkinsonian syndromes, clinically significant musculoskeletal or vestibular disorders affecting mobility, unstable antiparkinsonian therapy within the preceding three months, major visual impairment, or established cognitive impairment.

### Study procedures

Participants completed a series of assessments conducted in a single day. Evaluations were conducted in a prespecified sequence with rest intervals to mitigate fatigue. All clinical assessments were performed following routine dopaminergic administration, once participants reported experiencing their usual ON state. Where delayed ON occurred, assessment was postponed until the ON state had stabilised.

HRQoL, the primary outcome, was measured using the PDQ-39 Summary Index (PDQ-39 SI)[33]. The PDQ-39 comprises 39 items across eight domains scored on five-point Likert scales. Domain scores are transformed to a 0-100 scale, with higher scores denoting poorer HRQoL. The SI represents the mean of the eight domain scores. Motor complication severity was assessed using Part IV of the MDS-UPDRS[30], range 0-24, evaluating dyskinesias, motor fluctuations, and their functional impact. Higher scores indicate greater severity. In a subset of participants, objective motor data were collected using the Parkinson’s KinetiGraph^TM^ (PKG), which was worn continuously for seven days. PKG-derived measures included median Dyskinesia Score (DK_50), Fluctuation Dyskinesia Score (FDS), and percentage time immobile as an estimate of OFF time[34,35]. These measures were analysed as complementary markers of free-living motor burden (SUPPLEMENTARY MATERIAL B).

Illness perceptions were evaluated using Part 2 of the IPQ-R[24], encompassing perceived consequences, personal control, treatment control, chronic and cyclical timeline beliefs, illness coherence, and emotional representations. Symptom-related cognitive-behavioural responses were assessed using the CBRQ[25,26], generating subscales for symptom focusing, catastrophising, damage beliefs, fear-avoidance, embarrassment-avoidance, all-or-nothing behaviour, and avoidance/resting behaviour. Higher scores reflect stronger endorsement of each construct. Domain descriptions and conceptual distinctions are detailed in SUPPLEMENTARY MATERIALS C-E.

Cognitive function was measured using the Montreal Cognitive Assessment (MoCA)[36]. Anxiety and depressive symptoms were assessed using the Hospital Anxiety and Depression Scale (HADS-A and HADS-D, respectively)[37]. Additional demographic and clinical variables collected included age, sex, self-reported race/ethnicity[38], disease duration, Hoehn & Yahr stage, levodopa equivalent daily dose (LEDD), and MDS-UPDRS Parts I-III scores, covering non-motor experiences of daily living, motor experiences of daily living, and motor examination, respectively[30].

### Statistical analysis

All analyses were conducted using IBM SPSS Statistics for Windows, Version 29.0 (IBM Corp., Armonk, NY, USA). Scale scores were computed when fewer than 5% of items within a scale were missing. Otherwise, the scale score was treated as missing. Continuous variables are presented as mean (SD) or median (IQR), as appropriate depending on data distribution, and categorical variables as frequencies and percentages. Normality was assessed visually and using the Shapiro-Wilk test. Group differences were examined using independent-samples t tests or Mann-Whitney U tests for continuous variables and χ^2^ or Fisher’s exact tests for categorical variables, as appropriate. Associations between motor complication severity (MDS-UPDRS Part IV), IPQ-R domains, CBRQ subscales, and HRQoL (PDQ-39 SI) were examined using two-sided Pearson’s or Spearman’s rank correlation coefficients, depending on distribution. Partial Spearman correlations were computed using rank-transformed variables controlling for prespecified covariates. Hierarchical multiple linear regression was performed to examine whether psychological variables explained additional variance in HRQoL beyond demographic and clinical factors. Demographic and clinical variables (age, sex, disease duration, MDS-UPDRS Part III, LEDD, HADS-A, HADS-D, and MoCA) were entered in Block 1, motor complication severity (MDS-UPDRS Part IV) in Block 2, and selected IPQ-R and CBRQ domains in Block 3. Model fit was evaluated using R^2^ and change in R^2^ (ΔR^2^). Assumptions of linear regression were assessed through inspection of residual plots and variance inflation factors. To reduce the risk of overfitting, the number of predictors was limited relative to the sample size. In the wearable subgroup, convergent validity between PKG metrics and MDS-UPDRS Part IV was assessed using Spearman’s correlations, and exploratory analyses were repeated within this subgroup. All tests were two-sided with α=0.05. Given the exploratory nature of the study, no formal correction for multiple comparisons was applied. Interpretation focused on the strength and consistency of observed associations.

No formal *a priori* power calculation was performed. The study was embedded within the aforementioned clinical trial of an interventional device delivering vibrotactile stimulation which informed the sample size. Consequently, the measures are explorative and modelling was approached conservatively and findings interpreted with appropriate caution.

## RESULTS

### Cohort characteristics

Between 1 September 2024 and 28 February 2025, 60 people were screened; 58 met eligibility criteria and were enrolled (SUPPLEMENTARY MATERIAL F). Data completeness was 100% across study measures. Valid 7-day PKG recordings were obtained in 47 participants in accordance with the protocol. Baseline characteristics are summarised in TABLE 1. Median age was 68.0 years (IQR 62.0-75.8), with median symptom duration 4.5 years (2.0-8.0). The cohort was ethnically diverse, with 48.3% identifying as Asian, Black, or mixed ethnicity. Disease severity was consistent with moderate-stage PD (median Hoehn & Yahr 3.0; MDS-UPDRS Part IV 9.0). HRQoL demonstrated marked variability with median PDQ-39 SI 31.3 (17.7-43.5). In the PKG subset (n=47), demographic and clinical characteristics were comparable to the full cohort (SUPPLEMENTARY MATERIAL G: TABLE 5), with no significant differences in age (P=0.605), sex distribution (P=1.000), disease duration (P=0.170), MDS-UPDRS Part III (P=0.968), MDS-UPDRS Part IV (P=0.146), or PDQ-39 SI (P=0.362).

**Table 1.**
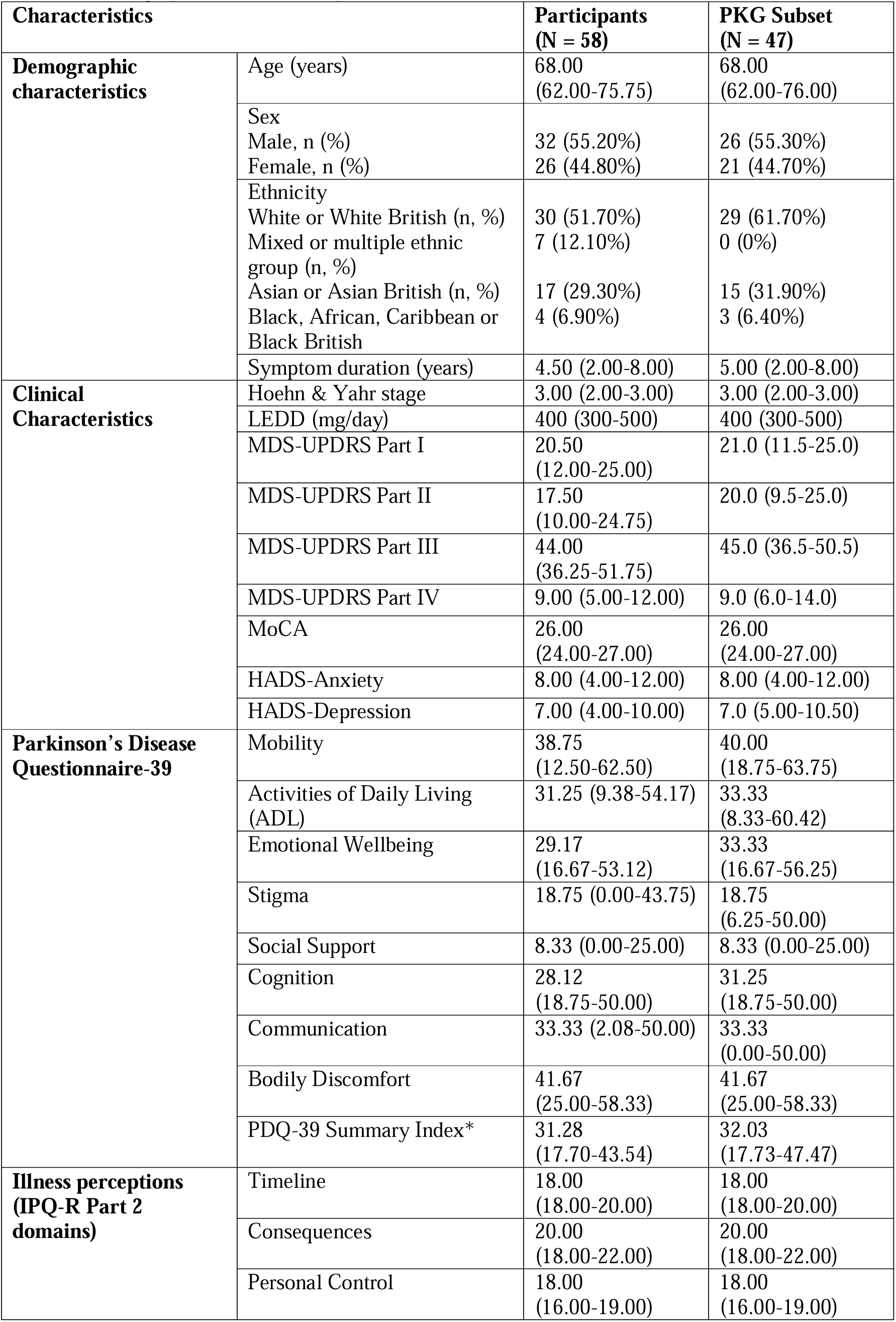
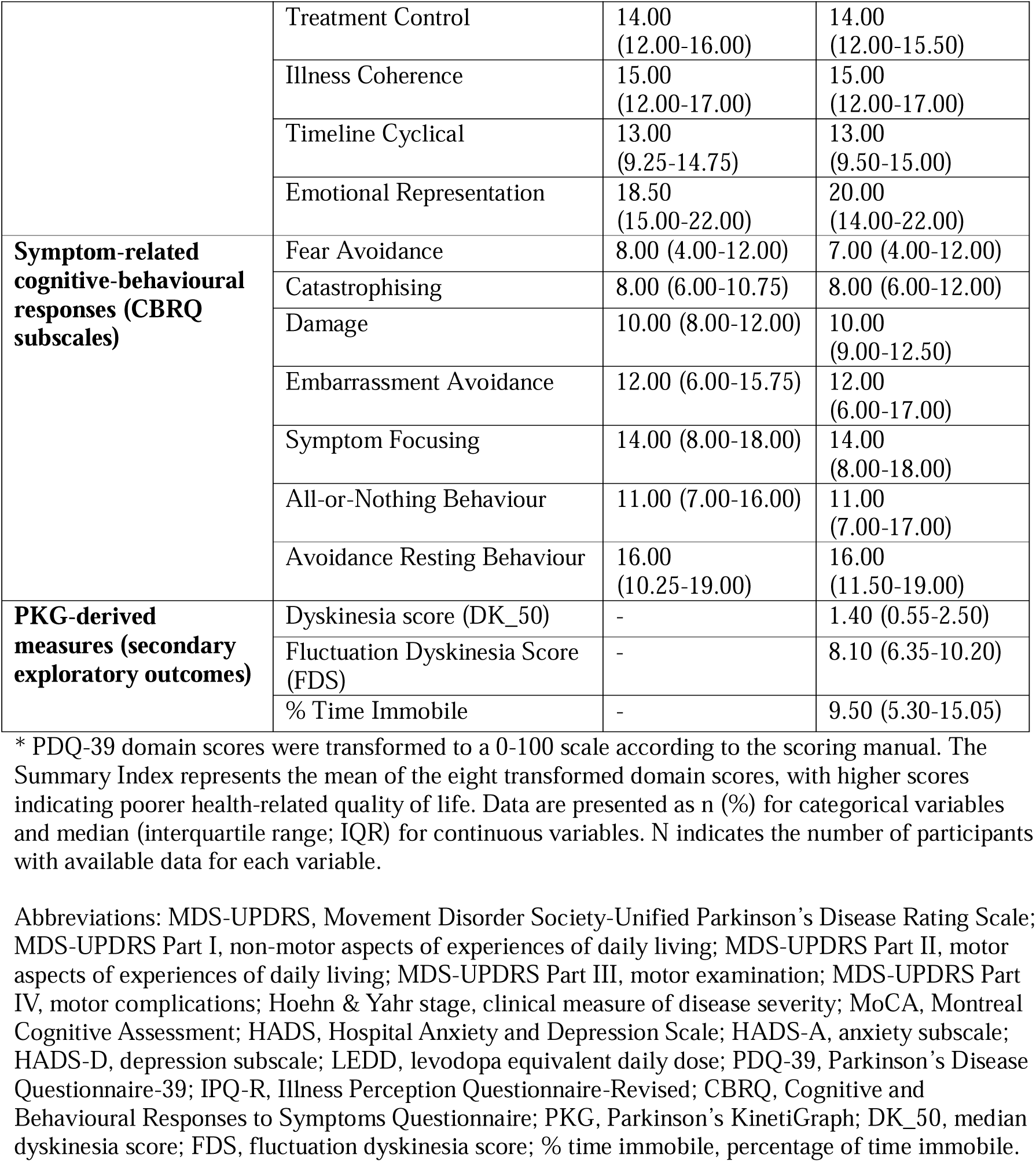
Demographic, clinical, and questionnaire characteristics.

### Motor complications and psychological constructs

Clinician-rated motor complication severity (MDS-UPDRS Part IV) demonstrated moderate associations with several symptom-level cognitive-behavioural responses (CBRQ domains), including catastrophising (ρ=0.482, P<0.001), symptom focusing (ρ=0.568, P<0.001), damage beliefs (ρ=0.404, P=0.002), avoidance/resting behaviour (ρ=0.458, P<0.001), and all-or-nothing behaviour (ρ=0.316, P=0.016) (TABLE 2). These domains reflect maladaptive interpretations of symptoms (e.g., exaggerated threat appraisal) and behavioural responses such as activity restriction or boom-bust patterns. Associations with illness-level beliefs (IPQ-R domains) were more selective, confined to cyclical timeline beliefs (ρ=0.426, P<0.001), reflecting perceived symptom variability and emotional representations (ρ=0.394, P=0.002), indicating illness-related distress. Other IPQ-R domains were not significantly related. Emotional representations showed a strong association with catastrophising (ρ=0.707, P<0.001) (SUPPLEMENTARY MATERIAL I), suggesting convergence between global emotional illness responses and symptom-specific threat appraisals.

**Table 2.**
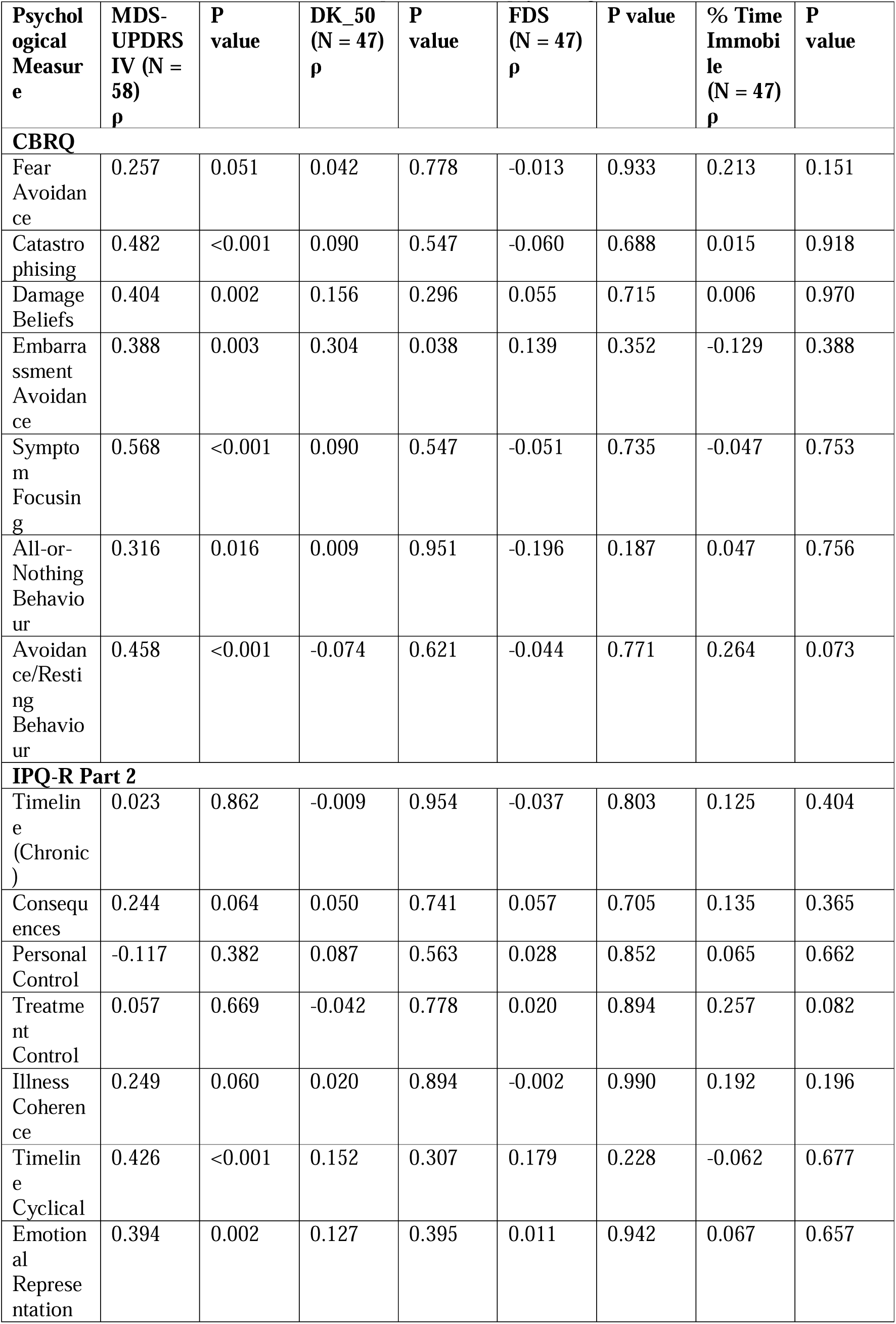
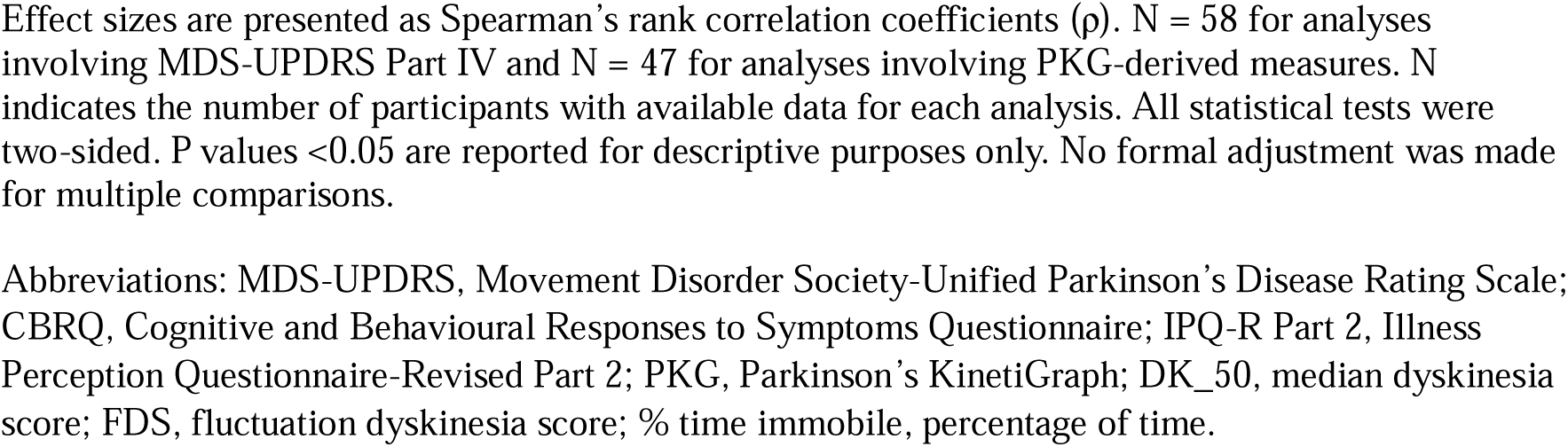
Associations between motor complications and psychological measures.

| Psychological Measure | MDS-UPDRS IV (N = 58)<br>$\rho$ | P value | DK_50 (N = 47)<br>$\rho$ | P value | FDS (N = 47)<br>$\rho$ | P value | % Time Immobile (N = 47)<br>$\rho$ | P value |
| --- | --- | --- | --- | --- | --- | --- | --- | --- |
| **CBRQ** |  |  |  |  |  |  |  |  |
| Fear Avoidance | 0.257 | 0.051 | 0.042 | 0.778 | -0.013 | 0.933 | 0.213 | 0.151 |
| Catastrophising | 0.482 | <0.001 | 0.090 | 0.547 | -0.060 | 0.688 | 0.015 | 0.918 |
| Damage Beliefs | 0.404 | 0.002 | 0.156 | 0.296 | 0.055 | 0.715 | 0.006 | 0.970 |
| Embarrassment Avoidance | 0.388 | 0.003 | 0.304 | 0.038 | 0.139 | 0.352 | -0.129 | 0.388 |
| Symptom Focusing | 0.568 | <0.001 | 0.090 | 0.547 | -0.051 | 0.735 | -0.047 | 0.753 |
| All-or-Nothing Behaviour | 0.316 | 0.016 | 0.009 | 0.951 | -0.196 | 0.187 | 0.047 | 0.756 |
| Avoidance/Resting Behaviour | 0.458 | <0.001 | -0.074 | 0.621 | -0.044 | 0.771 | 0.264 | 0.073 |
| **IPQ-R Part 2** |  |  |  |  |  |  |  |  |
| Timeline (Chronic) | 0.023 | 0.862 | -0.009 | 0.954 | -0.037 | 0.803 | 0.125 | 0.404 |
| Consequences | 0.244 | 0.064 | 0.050 | 0.741 | 0.057 | 0.705 | 0.135 | 0.365 |
| Personal Control | -0.117 | 0.382 | 0.087 | 0.563 | 0.028 | 0.852 | 0.065 | 0.662 |
| Treatment Control | 0.057 | 0.669 | -0.042 | 0.778 | 0.020 | 0.894 | 0.257 | 0.082 |
| Illness Coherence | 0.249 | 0.060 | 0.020 | 0.894 | -0.002 | 0.990 | 0.192 | 0.196 |
| Timeline Cyclical | 0.426 | <0.001 | 0.152 | 0.307 | 0.179 | 0.228 | -0.062 | 0.677 |
| Emotional Representation | 0.394 | 0.002 | 0.127 | 0.395 | 0.011 | 0.942 | 0.067 | 0.657 |

Effect sizes are presented as Spearman's rank correlation coefficients ( $\rho$ ). $N = 58$ for analyses involving MDS-UPDRS Part IV and $N = 47$ for analyses involving PKG-derived measures. $N$ indicates the number of participants with available data for each analysis. All statistical tests were two-sided. $P$ values $<0.05$ are reported for descriptive purposes only. No formal adjustment was made for multiple comparisons.
Abbreviations: MDS-UPDRS, Movement Disorder Society-Unified Parkinson's Disease Rating Scale; CBRQ, Cognitive and Behavioural Responses to Symptoms Questionnaire; IPQ-R Part 2, Illness Perception Questionnaire-Revised Part 2; PKG, Parkinson's KinetiGraph; DK\_50, median dyskinesia score; FDS, fluctuation dyskinesia score; % time immobile, percentage of time.

PKG-derived metrics demonstrated minimal associations with psychological measures; only embarrassment avoidance correlated with DK_50 (ρ=0.304, P=0.038). PKG measures were not significantly associated with MDS-UPDRS Part IV (SUPPLEMENTARY MATERIAL J). Participants with and without valid PKG data did not differ significantly (SUPPLEMENTARY MATERIAL G: TABLE 5).

### Determinants of HRQoL

Worse HRQoL correlated with greater motor complication severity (ρ=0.605, P<0.001) (TABLE 3). All CBRQ subscales were significantly associated with PDQ-39 SI (ρ=0.505-0.669; P<0.001). Among illness perceptions, emotional representations (ρ=0.702) and cyclical timeline beliefs (ρ=0.583) showed the strongest associations. After covariate adjustment, MDS-UPDRS Part IV remained independently associated with HRQoL (partial ρ=0.356, P=0.006) (SUPPLEMENTARY MATERIAL H). In the PKG subset (n=47), the pattern of associations was consistent (SUPPLEMENTARY MATERIAL H), with MDS-UPDRS Part IV remaining significantly associated with PDQ-39 SI (ρ=0.623, P<0.001). PKG-derived metrics (DK_50, FDS, % time immobile) were not significantly associated with HRQoL in adjusted or unadjusted analyses, although % time immobile showed a non-significant trend (ρ=0.271, P=0.067).

**Table 3.** Associations between clinical, psychological, and digital motor measures and health related quality of life.

| Variable | $\rho$ with PDQ-39 SI<br>(N=58) | P<br>value | $\rho$ with PDQ-39 SI<br>(N=47) | P<br>value |
| --- | --- | --- | --- | --- |
| **Motor complications (clinical)** |  |  |  |  |
| MDS-UPDRS Part IV | 0.605 | <0.001 | 0.641 | <0.001 |
| **Motor complications (PKG)** |  |  |  |  |
| DK_50 | - | - | 0.113 | 0.449 |
| FDS | - | - | 0.037 | 0.803 |
| % Time Immobile | - | - | 0.016 | 0.914 |
| **CBRQ subscales** |  |  |  |  |
| Fear Avoidance | 0.505 | <0.001 | 0.486 | 0.001 |
| Catastrophising | 0.669 | <0.001 | 0.676 | <0.001 |
| Damage Beliefs | 0.575 | <0.001 | 0.553 | <0.001 |
| Embarrassment Avoidance | 0.537 | <0.001 | 0.512 | <0.001 |
| Symptom Focusing | 0.581 | <0.001 | 0.573 | <0.001 |
| All-or-Nothing Behaviour | 0.610 | <0.001 | 0.601 | <0.001 |
| Avoidance/Resting Behaviour | 0.539 | <0.001 | 0.544 | <0.001 |
| **IPQ-R Part 2 domains** |  |  |  |  |
| Timeline (Chronic) | 0.154 | 0.249 | 0.163 | 0.275 |
| Consequences | 0.388 | 0.003 | 0.382 | 0.008 |
| Personal Control | -0.042 | 0.754 | -0.055 | 0.713 |
| Treatment Control | 0.315 | 0.016 | 0.311 | 0.034 |
| Illness Coherence | 0.336 | 0.010 | 0.341 | 0.020 |
| Timeline Cyclical | 0.583 | <0.001 | 0.580 | <0.001 |
| Emotional Representation | 0.702 | <0.001 | 0.712 | <0.001 |

Effect sizes are presented as Spearman's rank correlation coefficients ( $\rho$ ). N = 58 for the full cohort and N = 47 for the PKG subset; N indicates the number of participants with available data for each analysis. The PDQ-39 Summary Index (SI) was calculated as the mean of the eight transformed PDQ-39 domain scores (0-100 scale), with higher scores indicating poorer health-related quality of life. Dashes indicate variables not available in the full cohort. All statistical tests were two-sided. P values <0.05 are reported for descriptive purposes only. No formal adjustment was made for multiple comparisons.
Abbreviations: PDQ-39 SI, Parkinson's Disease Questionnaire-39 Summary Index; MDS-UPDRS, Movement Disorder Society-Unified Parkinson's Disease Rating Scale; CBRQ, Cognitive and Behavioural Responses to Symptoms Questionnaire; IPQ-R Part 2, Illness Perception Questionnaire-Revised Part 2; PKG, Parkinson's KinetiGraph; DK\_50, median dyskinesia score; FDS, fluctuation dyskinesia score; % time immobile, percentage of time immobile.

### Multivariate regression model of HRQoL

In hierarchical regression (TABLE 4), covariates explained 77.3% of HRQoL variance. Adding MDS-UPDRS Part IV increased explained variance (ΔR^2^=0.037, P=0.004). Inclusion of perceived consequences and catastrophising produced a further increase (ΔR^2^=0.041, P=0.004), yielding final R^2^=0.851 (adjusted R^2^=0.815). In the fully adjusted model, catastrophising (B=0.797, P=0.027) and perceived consequences (B=0.767, P=0.013) remained significant, whereas MDS-UPDRS Part IV did not (P=0.053). Collinearity diagnostics were acceptable. SUPPLEMENTARY MATERIAL G (TABLE 1) shows the incremental contribution of psychological variables, with IPQ-R consequences alone increasing R^2^ by 0.024 (P=0.011) and catastrophising alone by 0.020 (P=0.025), while their combined addition produced the greatest change (ΔR^2^=0.041, P=0.004). Collinearity diagnostics demonstrated acceptable variance inflation factors for all predictors (variance inflation factor range 1.35-2.57; SUPPLEMENTARY MATERIAL G: TABLE 2). Replacing DK_50 for MDS-UPDRS Part IV did not increase explained variance (SUPPLEMENTARY MATERIAL G: TABLE 6). Specifically, substituting DK_50 for MDS-UPDRS Part IV increased R² by only 0.013 (P=0.110), suggesting that objective dyskinesia severity did not make a significant contribution beyond the covariates (SUPPLEMENTARY MATERIAL G: TABLE 6). In the PKG subset hierarchical model (SUPPLEMENTARY MATERIAL G: TABLE 3), the final model explained 87.3% of variance (adjusted R^2^=0.833), with IPQ-R consequences remaining significant (B=0.785, P=0.028), whereas catastrophising and MDS-UPDRS Part IV were not independently significant (SUPPLEMENTARY MATERIAL G: TABLE 4).

**Table 4.**
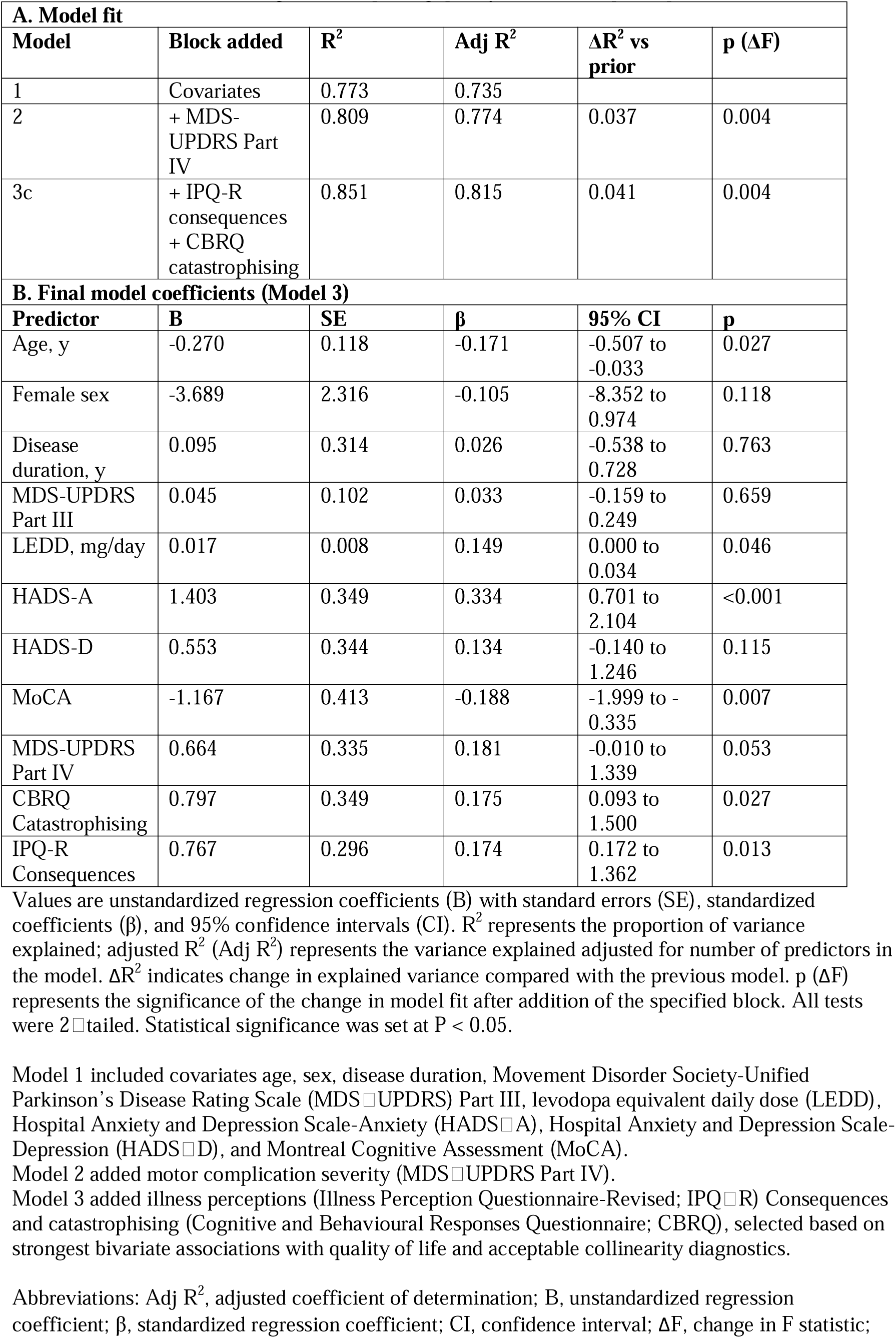
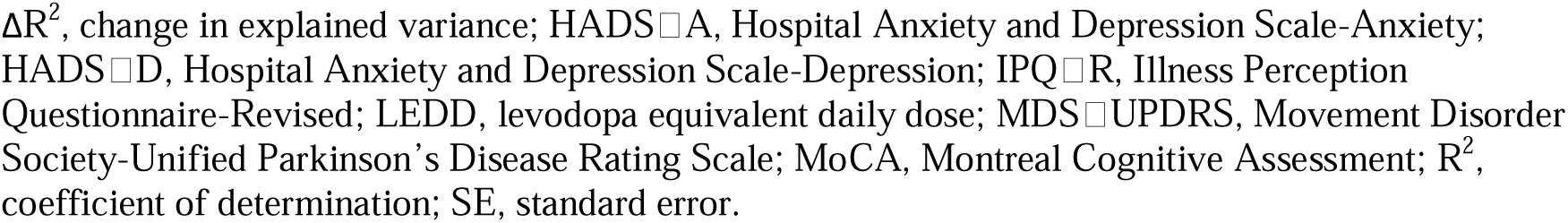
Hierarchical linear regression exploring quality of life in 58 participants.

| **A. Model fit** |  |  |  |  |  |
| --- | --- | --- | --- | --- | --- |
| **Model** | **Block added** | **R<sup>2</sup>** | **Adj R<sup>2</sup>** | **ΔR<sup>2</sup> vs prior** | **p (ΔF)** |
| 1 | Covariates | 0.773 | 0.735 |  |  |
| 2 | + MDS-UPDRS Part IV | 0.809 | 0.774 | 0.037 | 0.004 |
| 3c | + IPQ-R consequences<br>+ CBRQ catastrophising | 0.851 | 0.815 | 0.041 | 0.004 |
| **B. Final model coefficients (Model 3)** |  |  |  |  |  |
| **Predictor** | **B** | **SE** | **β** | **95% CI** | **p** |
| Age, y | -0.270 | 0.118 | -0.171 | -0.507 to -0.033 | 0.027 |
| Female sex | -3.689 | 2.316 | -0.105 | -8.352 to 0.974 | 0.118 |
| Disease duration, y | 0.095 | 0.314 | 0.026 | -0.538 to 0.728 | 0.763 |
| MDS-UPDRS Part III | 0.045 | 0.102 | 0.033 | -0.159 to 0.249 | 0.659 |
| LEDD, mg/day | 0.017 | 0.008 | 0.149 | 0.000 to 0.034 | 0.046 |
| HADS-A | 1.403 | 0.349 | 0.334 | 0.701 to 2.104 | <0.001 |
| HADS-D | 0.553 | 0.344 | 0.134 | -0.140 to 1.246 | 0.115 |
| MoCA | -1.167 | 0.413 | -0.188 | -1.999 to -0.335 | 0.007 |
| MDS-UPDRS Part IV | 0.664 | 0.335 | 0.181 | -0.010 to 1.339 | 0.053 |
| CBRQ Catastrophising | 0.797 | 0.349 | 0.175 | 0.093 to 1.500 | 0.027 |
| IPQ-R Consequences | 0.767 | 0.296 | 0.174 | 0.172 to 1.362 | 0.013 |

Values are unstandardized regression coefficients (B) with standard errors (SE), standardized coefficients (β), and 95% confidence intervals (CI). R<sup>2</sup> represents the proportion of variance explained; adjusted R<sup>2</sup> (Adj R<sup>2</sup>) represents the variance explained adjusted for number of predictors in the model. ΔR<sup>2</sup> indicates change in explained variance compared with the previous model. p (ΔF) represents the significance of the change in model fit after addition of the specified block. All tests were 2-tailed. Statistical significance was set at P < 0.05.
Model 1 included covariates age, sex, disease duration, Movement Disorder Society-Unified Parkinson's Disease Rating Scale (MDS-UPDRS) Part III, levodopa equivalent daily dose (LEDD), Hospital Anxiety and Depression Scale-Anxiety (HADS-A), Hospital Anxiety and Depression Scale-Depression (HADS-D), and Montreal Cognitive Assessment (MoCA).
Model 2 added motor complication severity (MDS-UPDRS Part IV).
Model 3 added illness perceptions (Illness Perception Questionnaire-Revised; IPQ-R) Consequences and catastrophising (Cognitive and Behavioural Responses Questionnaire; CBRQ), selected based on strongest bivariate associations with quality of life and acceptable collinearity diagnostics.
Abbreviations: Adj R<sup>2</sup>, adjusted coefficient of determination; B, unstandardized regression coefficient; β, standardized regression coefficient; CI, confidence interval; ΔF, change in F statistic;
$\Delta R^2$ , change in explained variance; HADS-A, Hospital Anxiety and Depression Scale-Anxiety; HADS-D, Hospital Anxiety and Depression Scale-Depression; IPQ-R, Illness Perception Questionnaire-Revised; LEDD, levodopa equivalent daily dose; MDS-UPDRS, Movement Disorder Society-Unified Parkinson's Disease Rating Scale; MoCA, Montreal Cognitive Assessment; $R^2$ , coefficient of determination; SE, standard error.

## DISCUSSION

This study examined whether psychological constructs contribute to HRQoL in PD beyond the measurable severity of motor complications. In these PD patients with moderate-stage disease, clinician-rated motor complication severity was strongly associated with HRQoL. However, when illness consequence beliefs and symptom-level catastrophising were introduced into multivariable models, both constructs accounted for additional variance, and the independent association between MDS-UPDRS Part IV and HRQoL was attenuated. These findings indicate that the perceived impact of motor instability is associated not only with its clinical severity but also with how it is interpreted (FIGURE 1).

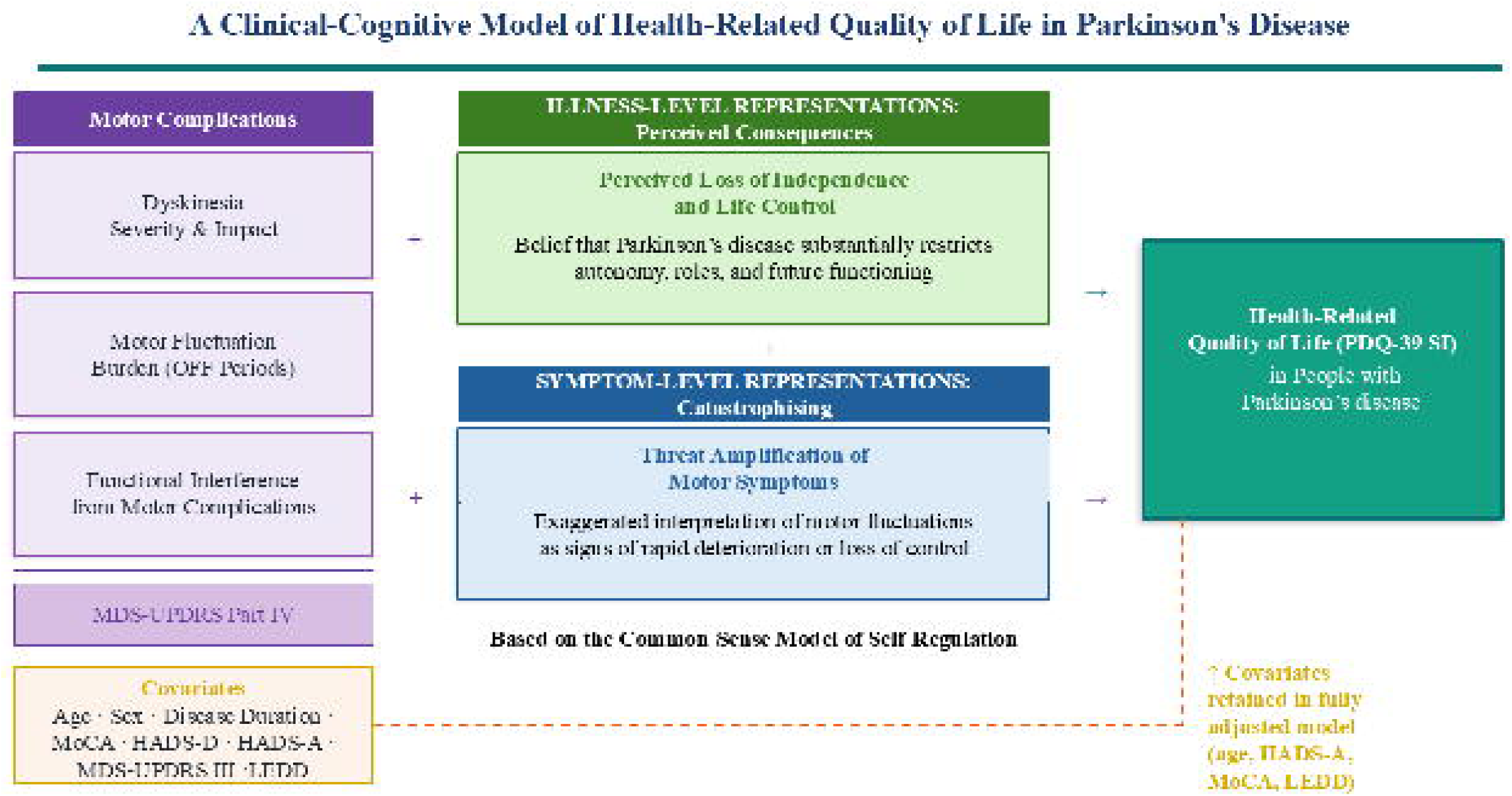

The relationship between motor complications and reduced HRQoL is well established[1,10,17,39]. Existing treatment pathways appropriately prioritise reduction of OFF time and troublesome dyskinesia[7], and improvements in motor stability are associated with gains in HRQoL[8,9]. Our data demonstrate that motor severity explains only part of the variability in patient-reported burden, consistent with prior observations that HRQoL is not fully determined by motor severity and complications[1,10,17,39].

The constructs that remained independently associated with HRQoL, perceived consequences and catastrophising, are conceptually consistent with the Common-Sense Model of self-regulation[21–23]. Consequence beliefs reflect broader evaluations regarding the extent to which PD disrupts autonomy and anticipated roles[24,40]. Catastrophising represents a proximal cognitive style characterised by threat amplification in response to symptom fluctuation[26]. Particularly, catastrophising retained statistical significance after adjustment for anxiety and depressive symptoms, aligning with evidence that distress during OFF periods and ruminative processes contribute independently to perceived burden[16,41,42]. Within this framework, people with comparable fluctuation profiles may differ in HRQoL according to interpretative stance rather than motor amplitude alone. Associations were observed between motor complication severity (e.g., dyskinesia, freezing, motor fluctuations) and behavioural responses such as avoidance and social withdrawal, with motor symptoms consistently linked to reduced participation[43]. Greater use of escape-avoidance coping predicted poorer mood and emotional well-being independent of disease severity, whereas planful problem-solving was associated with better communication and bodily discomfort outcomes[1]. Reduced motor autonomy was also linked to lower subjective and psychological well-being, including diminished life satisfaction and social contribution[13,14]. Additionally, social withdrawal in response to unpredictable or nonvisible symptoms was associated with decreased engagement and isolation[17,43]. These patterns may coexist with motor variability and contribute to perceived daily limitations beyond objective motor stage.

In contrast, wearable-derived PKG metrics were not associated with HRQoL and showed limited correspondence with clinician-rated motor complications. Although accelerometry validly quantifies bradykinesia, dyskinesia, and motor fluctuations in free-living environments[31,34,35] and can support therapeutic decision-making[2,7], it primarily captures motor kinematics rather than the perceived impact of symptoms on daily life. This distinction is clinically important. While sensor-based monitoring aims to reflect real-world motor behaviour, objective motor variability alone may be insufficient to explain patient-reported quality of life. MDS-UPDRS Part IV incorporates patient recall and functional consequence, thereby embedding subjective burden within the rating, whereas PKG metrics reflect continuous motor activity without reference to distress, interference, or meaning. Consistent with this, Moreau et al.[31] emphasise that wearable technologies complement but do not replace clinical evaluation, as they do not directly assess functional or psychosocial impact.

Strengths of this study include the concurrent modelling of clinician-rated motor complications, disease-specific HRQoL, validated assessment measures, and wearable motor quantification within a single analytic framework. Patients were recruited from a demographically diverse East London population, with substantial representation from minoritised racial and ethnic groups. This is methodologically important given documented disparities in HRQoL and cognitive outcomes across racial and ethnic groups in PD[27–29]. The present findings also support a self-regulatory model of HRQoL in PD, in which biological disease burden interacts with illness perceptions and cognitive-behavioural responses, while being shaped by the broader social and demographic context[21–24, 26–29]. Inclusion of a diverse clinical population therefore strengthens the external validity of these findings and supports their relevance to contemporary community PD care.

Several limitations warrant consideration. First, the cross-sectional design precludes conclusions regarding directionality or causal sequencing. It remains plausible that reduced HRQoL amplifies catastrophic interpretation, rather than catastrophic assessment contributing to diminished quality of life. Second, psychological constructs and HRQoL were assessed using self-report instruments, raising the possibility of shared-method variance influencing observed associations. Third, although models were specified conservatively, the modest sample size limits statistical power and may increase the risk of overfitting, particularly in multivariable analyses. Fourth, the wearable subset was smaller, potentially limiting detection of subtler associations between PKG-derived metrics and patient-reported outcomes. Finally, MDS-UPDRS Part IV incorporates patient recall and clinician judgement, and therefore is not a purely objective index of motor fluctuation and dyskinesia severity.

Motor optimisation remains central in PD management[2,7]. However, cognitive appraisal may meaningfully shape the perceived burden of motor complications. Hurt et al.[44] demonstrated that more negative illness perceptions, particularly stronger beliefs about consequences and greater symptom identity were associated with poorer HRQoL and mood, and that optimism moderated these relationships, buffering the impact of maladaptive beliefs. They also reported a non-linear association between optimism and mood, suggesting that adaptive, realistic positivity rather than excessive optimism is most beneficial[44]. Interventional evidence further supports clinical relevance. Cognitive Behavioural Therapy has been shown to reduce situational anxiety and avoidance behaviours in PD[45] and to produce significant reductions in anxiety and fear of falling[46], factors closely linked to motor presentation, activity restriction and social participation[43]. Future work should therefore examine whether targeting catastrophic interpretations of OFF periods and dyskinesia reduces perceived motor burden and improves HRQoL independently of objective motor severity.

## CONCLUSIONS

Motor complications remain central determinants of disability in PD. In this moderate-stage cohort, illness consequence beliefs and catastrophic interpretations of motor fluctuations and dyskinesias were independently associated with HRQoL beyond motor complication severity, mood, cognition, and dopaminergic exposure. Objective wearable-derived motor indices were not associated with HRQoL. These findings emphasise the potential value of integrating motor optimisation with structured assessment of illness beliefs in patient-centred care.

## Data Availability

De-identified participant data and supporting documents including study protocol, statistical analysis plan, data dictionary, and consent form will be made available after publication upon reasonable request to the corresponding author, subject to a data use agreement and applicable ethical constraints.

## ACKNOWLEDGMENTS

We thank all participants for taking part in this study. We also thank the study project team and supporting staff for their assistance. This work was supported by a UK Research and Innovation Knowledge Transfer Partnership (KTP) award (Innovate UK, 2021-2022, Round 4).

## REFERENCES

[1] Bucks, R. S., Cruise, K. E., Skinner, T. C., Loftus, A. M., Barker, R. A., & Thomas, M. G. (2011). Coping processes and health-related quality of life in Parkinson’s disease. International journal of geriatric psychiatry, 26(3), 247–255. 10.1002/gps.2520

[2] Bloem, B. R., Okun, M. S., & Klein, C. (2021). Parkinson’s disease. Lancet (London, England), 397(10291), 2284–2303. 10.1016/S0140-6736(21)00218-X

[3] Su, D., Cui, Y., He, C., Yin, P., Bai, R., Zhu, J., Lam, J. S. T., Zhang, J., Yan, R., Zheng, X., Wu, J., Zhao, D., Wang, A., Zhou, M., & Feng, T. (2025). Projections for prevalence of Parkinson’s disease and its driving factors in 195 countries and territories to 2050: modelling study of Global Burden of Disease Study 2021. BMJ (Clinical research ed.), 388, e080952. 10.1136/bmj-2024-080952

[4] Chang, H. J., Jang, M., Woo, K. A., Shin, J. H., Kim, H. J., & Jeon, B. (2024). The prevalence of non-troublesome dyskinesia in Parkinson’s disease. Parkinsonism & related disorders, 123, 106951. 10.1016/j.parkreldis.2024.106951

[5] di Biase, L., Pecoraro, P. M., Carbone, S. P., Caminiti, M. L., & Di Lazzaro, V. (2023). Levodopa-Induced Dyskinesias in Parkinson’s Disease: An Overview on Pathophysiology, Clinical Manifestations, Therapy Management Strategies and Future Directions. Journal of clinical medicine, 12(13), 4427. 10.3390/jcm12134427

[6] Kim, H. J., Mason, S., Foltynie, T., Winder-Rhodes, S., Barker, R. A., & Williams-Gray, C. H. (2020). Motor complications in Parkinson’s disease: 13-year follow-up of the CamPaIGN cohort. Movement disorders : official journal of the Movement Disorder Society, 35(1), 185–190. 10.1002/mds.27882

[7] de Bie, R. M. A., Katzenschlager, R., Swinnen, B. E. K. S., Peball, M., Lim, S. Y., Mestre, T. A., Perez Lloret, S., Coelho, M., Aquino, C., Tan, A. H., Bruno, V., Dijk, J. M., Heim, B., Lin, C. H., Kauppila, L. A., Litvan, I., Spijker, R., Seppi, K., Costa, J., Sampaio, C., … Silverdale, M. A. (2025). Update on Treatments for Parkinson’s Disease Motor Fluctuations - An International Parkinson and Movement Disorder Society Evidence-Based Medicine Review. Movement disorders : official journal of the Movement Disorder Society, 40(5), 776–794. 10.1002/mds.30162

[8] Santos-García, D., Solleiro, Á., González-Ortega, G., Mir, P., López-Ariztegui, N., Legarda, I., Rojas-Pérez, M. E., Peral-Quirós, A., Cabo, I., García-Ramos, R., Sánchez-Alonso, P., Hernández-Vara, J., Blázquez-Estrada, M., Sánchez-Ferro, Á., & DATs-PD GETM Spanish Registry Group (2025). Impact of device-aided therapies on quality of life in patients with Parkinson’s disease. A comparative multicenter observational study. Journal of neural transmission (Vienna, Austria : 1996), 10.1007/s00702-025-03066-z. Advance online publication. 10.1007/s00702-025-03066-z

[9] Ramirez-Zamora, A., Okun, M. S., Kukreja, P., & Hu, W. (2025). Parkinson’s disease quality of life at 12 months comparing invasive device-aided therapy with oral treatment. NPJ Parkinson’s disease, 11(1), 235. 10.1038/s41531-025-01093-x

[10] Schrag, A., Jahanshahi, M., & Quinn, N. (2000). What contributes to quality of life in patients with Parkinson’s disease?. Journal of neurology, neurosurgery, and psychiatry, 69(3), 308–312. 10.1136/jnnp.69.3.308

[11] Cilia, R., Akpalu, A., Sarfo, F. S., Cham, M., Amboni, M., Cereda, E., Fabbri, M., Adjei, P., Akassi, J., Bonetti, A., & Pezzoli, G. (2014). The modern pre-levodopa era of Parkinson’s disease: insights into motor complications from sub-Saharan Africa. Brain : a journal of neurology, 137(Pt 10), 2731–2742. 10.1093/brain/awu195

[12] Kelly, M. J., Lawton, M. A., Baig, F., Ruffmann, C., Barber, T. R., Lo, C., Klein, J. C., Ben-Shlomo, Y., & Hu, M. T. (2019). Predictors of motor complications in early Parkinson’s disease: A prospective cohort study. Movement disorders : official journal of the Movement Disorder Society, 34(8), 1174–1183. 10.1002/mds.27783

[13] Suzukamo, Y., Ohbu, S., Kondo, T., Kohmoto, J., & Fukuhara, S. (2006). Psychological adjustment has a greater effect on health-related quality of life than on severity of disease in Parkinson’s disease. Movement disorders : official journal of the Movement Disorder Society, 21(6), 761–766. 10.1002/mds.20817

[14] Vescovelli, F., Sarti, D., & Ruini, C. (2018). Subjective and psychological well-being in Parkinson’s Disease: A systematic review. Acta neurologica Scandinavica, 138(1), 12–23. 10.1111/ane.12946

[15] Heimrich, K. G., Schönenberg, A., Santos-García, D., Mir, P., Coppadis Study Group, & Prell, T. (2023). The Impact of Nonmotor Symptoms on Health-Related Quality of Life in Parkinson’s Disease: A Network Analysis Approach. Journal of clinical medicine, 12(7), 2573. 10.3390/jcm12072573

[16] Blundell, E. K., Grover, L. E., Stott, J., & Schrag, A. (2023). The experience of Anxiety for people with Parkinson’s disease. NPJ Parkinson’s disease, 9(1), 75. 10.1038/s41531-023-00512-1

[17] van Munster, M., Pedrosa, A. J., Künkler, C., & Pedrosa, D. J. (2024). The Quality in Quality of Life in Parkinson’s Disease: A Qualitative Meta-Synthesis. Movement disorders clinical practice, 11(7), 761–769. 10.1002/mdc3.14063

[18] Cassidy, I., Doody, O., Richardson, M., & Meskell, P. (2024). Quality of life and living with Parkinson’s disease: a qualitative exploration within an Irish context. BMC neurology, 24(1), 275. 10.1186/s12883-024-03769-y

[19] Smith, L. J., Callis, J., Bridger-Smart, S., & Guilfoyle, O. (2024). Experiences of Living With the Nonmotor Symptoms of Parkinson’s Disease: A Photovoice Study. Health expectations : an international journal of public participation in health care and health policy, 27(3), e14124. 10.1111/hex.14124

[20] Crooks, S., Mitchell, G., Wynne, L., & Carter, G. (2025). Exploring the stigma experienced by people affected by Parkinson’s disease: a systematic review. BMC public health, 25(1), 25. 10.1186/s12889-024-21236-8

[21] Leventhal H, Meyer D, Nerenz D. (1980). The common-sense representation of illness danger. In: Medical Psychology.

[22] Leventhal H, Brissette I, Leventhal EA. (2003). The common-sense model of self-regulation of health and illness.

[23] Hagger, M. S., & Orbell, S. (2022). The common sense model of illness self-regulation: a conceptual review and proposed extended model. Health psychology review, 16(3), 347–377. 10.1080/17437199.2021.1878050

[24] Moss-Morris R, Weinman J, Petrie KJ, Horne R, Cameron LD, Buick D. (2002). The Revised Illness Perception Questionnaire (IPQ-R). Psychol Health., 17(1), 1–16. 10.1080/08870440290001494

[25] Skerrett TN, Moss-Morris R. (2006). Fatigue and social impairment in multiple sclerosis: the role of patients’ cognitive and behavioral responses to their symptoms. J Psychosom Res, 61(5), 587–93. 10.1016/j.jpsychores.2006.04.018

[26] Picariello, F., Chilcot, J., Chalder, T., Herdman, D., & Moss-Morris, R. (2023). The Cognitive and Behavioural Responses to Symptoms Questionnaire (CBRQ): Development, reliability and validity across several long-term conditions. British journal of health psychology, 28(2), 619–638. 10.1111/bjhp.12644

[27] Di Luca, D. G., Luo, S., Liu, H., Cohn, M., Davis, T. L., Ramirez-Zamora, A., Rafferty, M., Dahodwala, N., Naito, A., Neault, M., Beck, J., & Marras, C. (2023). Racial and Ethnic Differences in Health-Related Quality of Life for Individuals With Parkinson Disease Across Centers of Excellence. Neurology, 100(21), e2170–e2181. 10.1212/WNL.0000000000207247

[28] Valenzuela, Y., Luna, K., Uribe-Kirby, R., Pawlak, A., Pitman, L., Cuellar-Rocha, P., Lucatero, G. R., Santos, M. M., & Jones, J. D. (2025). Cognitive Performance of Latino and White Non-Latino Individuals With Parkinson’s Disease. The Journal of neuropsychiatry and clinical neurosciences, 37(1), 14–19. 10.1176/appi.neuropsych.20240006

[29] Zirra, A., Dey, K. C., Camboe, E., Waters, S., Haque, T., Huxford, B., Chohan, H., Donkor, N., Kahan, J., Ben-Joseph, A., Gallagher, D. A., Budu, C., Boyle, T., Simonet, C., Lees, A. J., Marshall, C. R., & Noyce, A. J. (2025). The East London Parkinson’s disease project - a case-control study of Parkinson’s Disease in a diverse population. NPJ Parkinson’s disease, 11(1), 172. 10.1038/s41531-025-01031-x

[30] Goetz, C. G., Tilley, B. C., Shaftman, S. R., Stebbins, G. T., Fahn, S., Martinez-Martin, P., Poewe, W., Sampaio, C., Stern, M. B., Dodel, R., Dubois, B., Holloway, R., Jankovic, J., Kulisevsky, J., Lang, A. E., Lees, A., Leurgans, S., LeWitt, P. A., Nyenhuis, D., Olanow, C. W., … Movement Disorder Society UPDRS Revision Task Force (2008). Movement Disorder Society-sponsored revision of the Unified Parkinson’s Disease Rating Scale (MDS-UPDRS): scale presentation and clinimetric testing results. Movement disorders : official journal of the Movement Disorder Society, 23(15), 2129–2170. 10.1002/mds.22340

[31] Moreau, C., Rouaud, T., Grabli, D., Benatru, I., Remy, P., Marques, A. R., Drapier, S., Mariani, L. L., Roze, E., Devos, D., Dupont, G., Bereau, M., & Fabbri, M. (2023). Overview on wearable sensors for the management of Parkinson’s disease. NPJ Parkinson’s disease, 9(1), 153. 10.1038/s41531-023-00585-y

[32] Postuma, R. B., Berg, D., Stern, M., Poewe, W., Olanow, C. W., Oertel, W., Obeso, J., Marek, K., Litvan, I., Lang, A. E., Halliday, G., Goetz, C. G., Gasser, T., Dubois, B., Chan, P., Bloem, B. R., Adler, C. H., & Deuschl, G. (2015). MDS clinical diagnostic criteria for Parkinson’s disease. Movement disorders : official journal of the Movement Disorder Society, 30(12), 1591–1601. 10.1002/mds.26424

[33] Jenkinson, C., Fitzpatrick, R., Peto, V., Greenhall, R., & Hyman, N. (1997). The Parkinson’s Disease Questionnaire (PDQ-39): development and validation of a Parkinson’s disease summary index score. Age and ageing, 26(5), 353–357. 10.1093/ageing/26.5.353

[34] Griffiths, R. I., Kotschet, K., Arfon, S., Xu, Z. M., Johnson, W., Drago, J., Evans, A., Kempster, P., Raghav, S., & Horne, M. K. (2012). Automated assessment of bradykinesia and dyskinesia in Parkinson’s disease. Journal of Parkinson’s disease, 2(1), 47–55. 10.3233/JPD-2012-11071

[35] Farzanehfar, P., Woodrow, H., Braybrook, M., McGregor, S., Evans, A., Nicklason, F., & Horne, M. (2018). Objective measurement in routine care of people with Parkinson’s disease improves outcomes. NPJ Parkinson’s disease, 4, 10. 10.1038/s41531-018-0046-4

[36] Hoops, S., Nazem, S., Siderowf, A. D., Duda, J. E., Xie, S. X., Stern, M. B., & Weintraub, D. (2009). Validity of the MoCA and MMSE in the detection of MCI and dementia in Parkinson disease. Neurology, 73(21), 1738–1745. 10.1212/WNL.0b013e3181c34b47

[37] Zigmond AS, Snaith RP. (1983). The hospital anxiety and depression scale. Acta Psychiatrica Scandinavica, 67(6), 361–70. 10.1111/j.1600-0447.1983.tb09716.x.

[38] Office for National Statistics (ONS). (2021).Ethnic group, England and Wales: Census 2021 (classification and guidance). Available from: https://www.ons.gov.uk/

[39] Hechtner, M. C., Vogt, T., Zöllner, Y., Schröder, S., Sauer, J. B., Binder, H., Singer, S., & Mikolajczyk, R. (2014). Quality of life in Parkinson’s disease patients with motor fluctuations and dyskinesias in five European countries. Parkinsonism & related disorders, 20(9), 969–974. 10.1016/j.parkreldis.2014.06.001

[40] Evans, D., & Norman, P. (2009). Illness representations, coping and psychological adjustment to Parkinson’s disease. Psychology & health, 24(10), 1181–1196. 10.1080/08870440802398188

[41] Fernie, B. A., Spada, M. M., & Brown, R. G. (2019). Motor fluctuations and psychological distress in Parkinson’s disease. Health psychology : official journal of the Division of Health Psychology, American Psychological Association, 38(6), 518–526. 10.1037/hea0000736

[42] Julien, C. L., Rimes, K. A., & Brown, R. G. (2016). Rumination and behavioural factors in Parkinson’s disease depression. Journal of psychosomatic research, 82, 48–53. 10.1016/j.jpsychores.2016.01.008

[43] Ahn, S., Springer, K., & Gibson, J. S. (2022). Social withdrawal in Parkinson’s disease: A scoping review. Geriatric nursing (New York, N.Y.), 48, 258–268. 10.1016/j.gerinurse.2022.10.010

[44] Hurt, C. S., Burn, D. J., Hindle, J., Samuel, M., Wilson, K., & Brown, R. G. (2014). Thinking positively about chronic illness: An exploration of optimism, illness perceptions and well-being in patients with Parkinson’s disease. British journal of health psychology, 19(2), 363–379. 10.1111/bjhp.12043

[45] Mulders, A. E. P., Moonen, A. J. H., Dujardin, K., Kuijf, M. L., Duits, A., Flinois, B., Handels, R. L. H., Lopes, R., & Leentjens, A. F. G. (2018). Cognitive behavioural therapy for anxiety disorders in Parkinson’s disease: Design of a randomised controlled trial to assess clinical effectiveness and changes in cerebral connectivity. Journal of psychosomatic research, 112, 32–39. 10.1016/j.jpsychores.2018.04.002

[46] Reynolds, G. O., Saint-Hilaire, M., Thomas, C. A., Barlow, D. H., & Cronin-Golomb, A. (2020). Cognitive-Behavioral Therapy for Anxiety in Parkinson’s Disease. Behavior modification, 44(4), 552–579. 10.1177/0145445519838828

