## Supplemental Files for "Perceived Consequences and Catastrophising Help Explain Health-Related Quality of Life in Parkinson’s Disease. A Cross-Sectional Study"

### **Supplementary File**

#### **Supplementary material A.** STROBE Statement-Checklist of items that should be included in reports of cross-sectional studies

|  | **Item No** | **Recommendation** | **Reported on page/section** |
| --- | --- | --- | --- |
| **Title and abstract** | 1 | (a) Indicate the study’s design with a commonly used term in the title or the abstract | Title page,  abstract (p.2-3) |
|  |  | (b) Provide in the abstract an informative and balanced summary of what was done and what was found | Abstract (p.2-3) |
| **Introduction** | | |  |
| Background/rationale | 2 | Explain the scientific background and rationale for the investigation being reported | Introduction (p.3-6) |
| Objectives | 3 | State specific objectives, including any prespecified hypotheses | End of introduction, Aims (p.6) |
| **Methods** | | |  |
| Study design | 4 | Present key elements of study design early in the paper | Methods,  Study design (p.7) |
| Setting | 5 | Describe the setting, locations, and relevant dates, including periods of recruitment, exposure, follow-up, and data collection | Methods,  Participants (p.7),  Results (p.10-13) |
| Participants | 6 | (a) Give the eligibility criteria, and the sources and methods of selection of participants | Methods,  Participants (p.7) |
| Variables | 7 | Clearly define all outcomes, exposures, predictors, potential confounders, and effect modifiers. Give diagnostic criteria, if applicable | Methods,  Study procedures (p.8-9) |
| Data sources/ measurement | 8* | For each variable of interest, give sources of data and details of methods of assessment (measurement). Describe comparability of assessment methods if there is more than one group | Methods,  Study procedures (p.8-9), Supplementary materials B-E |
| Bias | 9 | Describe any efforts to address potential sources of bias | Methods,  Study procedures (p.8-9), statistical analysis (p.9-10) |
| Study size | 10 | Explain how the study size was arrived at | Methods, Sample size justification (p.10) |
| Quantitative variables | 11 | Explain how quantitative variables were handled in the analyses. If applicable, describe which groupings were chosen and why | Methods, Statistical analysis (p.9-10) |
| Statistical methods | 12 | (a) Describe all statistical methods, including those used to control for confounding | Methods, Statistical analysis (p.9-10) |
|  |  | (b) Describe any methods used to examine subgroups and interactions | Methods, Statistical analysis (p. 9-10) |
|  |  | (c) Explain how missing data were addressed | Methods, statistical analysis (p.9-10) |
|  |  | (d) If applicable, describe analytical methods taking account of sampling strategy | Not applicable (consecutive clinic sample) |
|  |  | (e) Describe any sensitivity analyses | Methods, statistical analysis (p.9-10), Supplementary material C |
| **Results** | | |  |
| Participants | 13* | (a) Report numbers of individuals at each stage of study—eg numbers potentially eligible, examined for eligibility, confirmed eligible, included in the study, completing follow-up, and analysed | Results on Participants (p.10-11), Supplementary material F |
|  |  | (b) Give reasons for non-participation at each stage | Results on Participants (p.10-11), Supplementary material F |
|  |  | (c) Consider use of a flow diagram | Supplementary material F |
| Descriptive data | 14* | (a) Give characteristics of study participants (eg demographic, clinical, social) and information on exposures and potential confounders | Results on Cohort characteristics (p.10-11), Table 1 |
|  |  | (b) Indicate number of participants with missing data for each variable of interest | Results on Cohort characteristics (p.10-11), Tables 1 |
| Outcome data | 15* | Report numbers of outcome events or summary measures | Results, Tables 2-4 and Supplementary materials F-I |
| Main results | 16 | (a) Give unadjusted estimates and, if applicable, confounder-adjusted estimates and their precision (eg, 95% confidence interval). Make clear which confounders were adjusted for and why they were included | Results, Tables 2-4 and Supplementary materials G-J |
|  |  | (b) Report category boundaries when continuous variables were categorized | Not applicable (no categorisation) |
|  |  | (c) If relevant, consider translating estimates of relative risk into absolute risk for a meaningful time period | Not applicable (continuous outcome) |
| Other analyses | 17 | Report other analyses done—eg analyses of subgroups and interactions, and sensitivity analyses | Results, (p.11-13),  Supplementary materials G-J |
| **Discussion** | | |  |
| Key results | 18 | Summarise key results with reference to study objectives | Discussion opening (p.13) |
| Limitations | 19 | Discuss limitations of the study, taking into account sources of potential bias or imprecision. Discuss both direction and magnitude of any potential bias | Discussion, Limitations (p.13-17) |
| Interpretation | 20 | Give a cautious overall interpretation of results considering objectives, limitations, multiplicity of analyses, results from similar studies, and other relevant evidence | Discussion, Relation to prior work, Possible mechanisms, Clinical implications (p.13-17) |
| Generalisability | 21 | Discuss the generalisability (external validity) of the study results | Discussion, Limitations (p.15-16) |
| **Other information** | | |  |
| Funding | 22 | Give the source of funding and the role of the funders for the present study and, if applicable, for the original study on which the present article is based | Title page (p.1), Funding Sources and Role of the Funder/Sponsors (p.17) |

*Give information separately for exposed and unexposed groups.

**Note:** An Explanation and Elaboration article discusses each checklist item and gives methodological background and published examples of transparent reporting. The STROBE checklist is best used in conjunction with this article (freely available on the Web sites of PLoS Medicine at http://www.plosmedicine.org/, Annals of Internal Medicine at http://www.annals.org/, and Epidemiology at http://www.epidem.com/). Information on the STROBE Initiative is available at [www.strobe-statement.org](http://www.strobe-statement.org).

#### **Supplementary material B.** Parkinson’s KinetiGraph measures, definitions and explanations

| **PKG measure** | **Definition** | **Explanation** |
| --- | --- | --- |
| Dyskinesia Score (DK_50) | Median dyskinesia score | A continuous measure derived from wrist accelerometry representing the median level of dyskinesia across the recording period. Higher values reflect more severe dyskinetic movements. |
| Fluctuation Dyskinesia Score (FDS) | Composite fluctuation index | A PKG-derived composite index that quantifies motor fluctuation by integrating variability in bradykinesia and dyskinesia over time. Higher values indicate greater fluctuation severity, consistent with OFF-ON oscillations. |
| Percentage Time Immobile | % of epochs with minimal movement | The percentage of time during which the wrist accelerometer detects minimal movement, interpreted as periods of immobility. It has been used as an objective proxy for OFF time or reduced mobility, particularly when assessed during typical waking hours. |

Abbreviations: PKG, Parkinson’s KinetiGraph; DK_50, median dyskinesia score; FDS, fluctuation dyskinesia score.

#### **Supplementary material C.** Contextual explanation of illness perceptions (Table A) and cognitive beliefs about symptoms (Table B) domains including explanation in relation to Parkinson’s disease

**Table A** Contextual explanation of illness perceptions (Illness Perception Questionnaire-Revised; IPQ-R) in relation to Parkinson’s disease

| **Domain** | **What It Measures** | **What It Might Mean in a Person with Parkinson’s** | **Example in Parkinson’s Context** | **Scoring System** |
| --- | --- | --- | --- | --- |
| **Identity** | Number of symptoms attributed to the illness | How many symptoms the person believes are caused by Parkinson’s disease | A person attributes tremor, stiffness, motor complications, fatigue, sleep problems, and mood changes to Parkinson’s disease | Count of symptoms endorsed as related to illness (Yes/No format). Higher score = stronger illness identity (more symptoms attributed to Parkinson’s disease) |
| **Timeline (Acute/Chronic)** | Beliefs about illness duration | Belief that Parkinson’s disease is permanent and progressive | ‘I will have Parkinson’s for the rest of my life.’ | 5-point Likert (Strongly Disagree-Strongly agree). Higher score = stronger belief that Parkinson’s disease is long-term |
| **Timeline (Cyclical)** | Beliefs about variability of symptoms | Recognition of delayed on, ‘on/off’ periods, wearing off, and dyskinesias | ‘My symptoms come and go in cycles.’ | 5-point Likert. Higher score = belief Parkinson’s disease symptoms are unpredictable/variable |
| **Consequences** | Perceived impact on life | Belief that Parkinson’s disease seriously affects independence, work, social roles | ‘Parkinson’s has major consequences on my life.’ | 5-point Likert. Higher score = perceived severe impact |
| **Personal Control** | Belief in own ability to influence illness | Belief that exercise, medication adherence, or stress management helps control symptoms | ‘What I do can influence my symptoms.’ | 5-point Likert. Higher score = greater perceived self-control |
| **Treatment Control** | Belief in treatment effectiveness | Confidence that levodopa or deep brain stimulation improves symptoms | ‘My treatment can control my illness.’ | 5-point Likert. Higher score = stronger belief in treatment effectiveness |
| **Illness Coherence** | Degree of understanding of illness | How well the person feels they understand Parkinson’s disease | ‘I have a clear understanding of my condition.’ | 5-point Likert. Higher score = better illness understanding |
| **Emotional Representations** | Emotional response to illness | Emotional distress related to Parkinson’s diagnosis or progression | ‘Thinking about my illness makes me anxious.’ | 5-point Likert. Higher score = greater emotional distress related to PD |

**Table B** Contextual explanation of Cognitive and Behavioural Responses to Symptoms Questionnaire (CBRQ) in relation to Parkinson’s disease

| **Domain** | **What It Measures** | **What It Might Mean in Parkinson’s** | **Example in Parkinson’s Context** | **Scoring System** |
| --- | --- | --- | --- | --- |
| **Fear Avoidance Beliefs** | Belief that activity worsens symptoms | Fear that walking or exercising will worsen tremor or cause falls | ‘If I move too much, my symptoms will get worse.’ | 0-4 Likert (Strongly Disagree-Strongly agree). Higher score = stronger fear-based beliefs |
| **Damage Beliefs** | Belief symptoms signal bodily harm | Interpreting stiffness or tremor as sign of brain deterioration | ‘My symptoms mean my body is being damaged.’ | 0-4 Likert. Higher score = stronger damage beliefs |
| **Catastrophising** | Exaggerated negative interpretation | Viewing minor symptom fluctuation as major decline | ‘This tremor means I’m getting rapidly worse.’ | 0-4 Likert. Higher score = greater catastrophising |
| **Embarrassment Avoidance** | Avoidance due to social concerns | Avoiding social situations due to visible tremor | ‘I avoid going out because people may notice my symptoms.’ | 0-4 Likert. Higher score = more socially driven avoidance |
| **Symptom Focusing** | Excessive attention to symptoms | Constant monitoring of tremor or gait | Frequently checking if hands are shaking | 0-4 Likert. Higher score = increased symptom monitoring |
| **Avoidance/Resting Behaviour** | Reducing activity due to symptoms | Staying seated most of the day to prevent fatigue or stiffness | Avoiding exercise to prevent worsening symptoms | 0-4 (Never-All the time). Higher score = greater inactivity |
| **All-or-Nothing Behaviour** | Boom-bust activity cycle | Overexerting on ‘good days,’ then severe fatigue on following days | Doing too much on medication ‘on’ periods then crashing | 0-4 (Never-All the time). Higher score = stronger boom-bust pattern |

#### **Supplementary material D.** General differences between the Illness Perception Questionnaire-Revised and the Cognitive and Behavioural Responses to Symptoms Questionnaire (Table A), and their relevance to Parkinson’s disease (Table B).

**Table A** General differences between the Illness Perception Questionnaire-Revised and the Cognitive and Behavioural Responses to Symptoms Questionnaire.

| **Dimension** | **Illness Perception Questionnaire- Revised** | **Cognitive and Behavioural Responses to Symptoms Questionnaire** |
| --- | --- | --- |
| **Theoretical Foundation** | Based on the Common-Sense Model of Self-Regulation. Proposes that individuals form structured cognitive and emotional representations of illness that guide coping and adjustment. | Based on cognitive behavioural theory and fear-avoidance models. Proposes that interpretations of symptoms influence behavioural patterns that may maintain or worsen disability. |
| **Core Conceptual Focus** | Global illness representations (what the illness means to the individual). | Cognitive and behavioural responses to specific symptoms (how the person reacts to symptom experiences). |
| **Level of Analysis** | Illness-level (macro cognitive framework). | Symptom-level (micro behavioural process). |
| **Primary Constructs Measured** | Identity, timeline beliefs, consequences, personal control, treatment control, illness coherence, emotional representations, and causal attributions. | Fear avoidance beliefs, damage beliefs, catastrophising, embarrassment avoidance, symptom focusing, avoidance or resting behaviour, and all-or-nothing behaviour. |
| **Nature of Constructs** | Primarily belief-based cognitive schemas about illness. | Combination of cognitive appraisals and observable behavioural responses. |
| **Emotional Measurement** | Directly measures emotional responses toward the illness itself. | Measures emotional processes indirectly through symptom-related fear and catastrophic interpretations. |
| **Behavioural Measurement** | Does not directly assess behaviour. Behaviour is inferred through coping theory. | Directly assesses behavioural responses such as activity avoidance, excessive resting, and boom–bust cycles. |
| **Temporal Orientation** | Long-term beliefs about illness duration, progression, and consequences. | Immediate and ongoing responses to current symptom experiences. |
| **Causal Attributions** | Explicitly measures perceived causes of illness (biological, psychological, environmental). | Does not assess illness causation. Focuses on reactions to symptoms rather than causes. |
| **Illness Understanding** | Includes a coherence dimension assessing how well the person understands their illness. | Does not assess overall illness understanding. |
| **Mechanism of Influence** | Beliefs influence coping strategies, emotional adjustment, adherence, and health behaviour. | Symptom interpretations influence behavioural responses, which may directly maintain or amplify disability. |
| **Intervention Sensitivity** | May change following psychoeducation or cognitive reframing of illness beliefs. | Particularly sensitive to cognitive behavioural therapy targeting maladaptive behaviours and symptom interpretations. |
| **Predictive Utility** | Commonly predicts treatment adherence, psychological adjustment, and quality of life. | Commonly predicts functional impairment, activity limitation, and persistence of symptoms. |
| **Scope of Application** | Broadly applicable across acute and chronic illnesses. | Particularly relevant in persistent symptom conditions where behaviour influences disability. |
| **Hierarchy of Processes** | Operates at a higher-order belief level influencing coping selection. | Operates at a lower-order process level influencing day-to-day functioning. |
| **Conceptual Strength** | Strong theoretical grounding in illness representation research. | Strong behavioural and mechanistic relevance to intervention and rehabilitation research. |
| **Primary Clinical Use** | Understanding illness meaning, acceptance, and treatment beliefs. | Identifying modifiable behavioural patterns that may sustain symptom-related disability. |
| **Key Conceptual Distinction** | Explains how a person conceptualizes their illness. | Explains how a person reacts to and behaves in response to symptoms. |

**Table B** Conceptual distinction between the Illness Perception Questionnaire-Revised and the Cognitive and Behavioural Responses to Symptoms Questionnaire in Parkinson’s disease

| **Analytical Dimension** | **Illness Perception Questionnaire-Revised** | **Cognitive and Behavioural Responses to Symptoms Questionnaire** |
| --- | --- | --- |
| **Theoretical Basis** | Grounded in the Common-Sense Model of Self-Regulation, which proposes that individuals construct cognitive and emotional representations of their illness that guide coping and adjustment. In Parkinson’s disease, this means the person develops beliefs about what the illness is, how long it will last, how serious it is, and whether it can be controlled. | Grounded in cognitive behavioural theory and fear-avoidance models of persistent symptoms. It focuses on how interpretations of specific symptoms (for example tremor, stiffness, fatigue, freezing of gait) influence behavioural responses. In Parkinson’s disease, this means examining how the person responds to symptoms in daily life rather than how they conceptualize the illness overall. |
| **Primary Focus** | Examines the person’s overall beliefs about Parkinson’s disease as a condition. It captures how they understand the illness, its causes, its impact, and its controllability. | Examines moment-to-moment cognitive and behavioural reactions to individual symptoms. It captures how the person interprets and reacts to tremor, fatigue, rigidity, or mobility changes in daily life. |
| **Level of Analysis** | Operates at an illness-level perspective. It evaluates how the person views Parkinson’s disease as a whole. | Operates at a symptom-level perspective. It evaluates how the person responds to specific symptom experiences within daily routines. |
| **Temporal Orientation** | Focuses on long-term beliefs about duration, progression, and overall consequences. For example, whether the person believes Parkinson’s disease is inevitably progressive and disabling. | Focuses on immediate or short-term interpretations and reactions. For example, whether a person avoids walking today because they fear falling due to current stiffness. |
| **Emotional Component** | Directly assesses emotional responses to Parkinson’s disease, such as fear, anxiety, anger, or distress when thinking about the illness. | Assesses emotional processes indirectly through beliefs such as catastrophising or fear of symptom worsening. The emotional component is embedded in how symptoms are interpreted rather than in global illness-related emotions. |
| **Behavioural Measurement** | Does not directly measure behavioural patterns. It assumes behaviour is influenced by beliefs but does not assess activity avoidance or overexertion. | Explicitly measures behavioural responses such as avoidance of movement, excessive resting, social withdrawal, or all-or-nothing activity patterns. In Parkinson’s disease, this might include staying seated to prevent tremor or overexerting on a ‘good’ medication day. |
| **Mechanism of Action** | Influences how the person selects coping strategies. For example, believing Parkinson’s disease is uncontrollable may reduce motivation to exercise or adhere to therapy. | Identifies mechanisms that may maintain or worsen disability. For example, avoiding walking due to fear of falling may lead to deconditioning, which then increases mobility limitations. |
| **Relevance to Parkinson’s Motor Symptoms** | Helps understand how a person interprets tremor, rigidity, or bradykinesia within the broader illness framework. For example, whether they see tremor as manageable or as a sign of inevitable decline. | Helps understand how the person reacts behaviourally to motor symptoms. For example, whether they avoid public spaces due to visible tremor or restrict activity due to stiffness. |
| **Relevance to Parkinson’s Non-Motor Symptoms** | Captures beliefs about fatigue, sleep disturbance, mood changes, or cognitive slowing as part of the illness identity and perceived consequences. | Captures behavioural responses to fatigue or cognitive slowing, such as resting excessively, withdrawing socially, or pushing through fatigue and then experiencing exhaustion. |
| **Use in Intervention Studies** | Useful for understanding illness acceptance, treatment beliefs, and psychological adjustment. It may predict adherence to medication or engagement in physiotherapy. | Particularly useful for identifying modifiable behavioural targets in psychological or rehabilitation interventions. It is sensitive to changes following cognitive behavioural therapy or structured activity programmes. |
| **Clinical Interpretation in Parkinson’s Disease** | A high score on chronic timeline and consequences suggests that the person perceives Parkinson’s disease as severely disabling and permanent. A low personal control score may indicate reduced self-efficacy. | High fear avoidance or avoidance behaviour scores suggest risk of reduced mobility and social withdrawal. High all-or-nothing behaviour may indicate fluctuating activity patterns that worsen fatigue. |
| **Clinical Implication** | Supports understanding of how a person cognitively and emotionally integrates Parkinson’s disease into their identity and life narrative. | Supports identification of behavioural patterns that may be maintaining disability and can be directly targeted in rehabilitation or psychological treatment. |

#### **Supplementary material E.** Relationship between illness perceptions, symptom responses, and quality of life in people with Parkinson’s disease

| **Quality of Life Domain in Parkinson’s Disease** | **Illness Perception Questionnaire-Revised: Influence on Quality of Life** | **Cognitive and Behavioural Responses to Symptoms Questionnaire: Influence on Quality of Life** |
| --- | --- | --- |
| **Overall Quality of Life** | Global beliefs about Parkinson’s disease (e.g., perceived severity, chronicity, and controllability) shape overall life satisfaction and illness acceptance. | Daily responses to tremor, rigidity, fatigue, and freezing of gait influence how much symptoms interfere with meaningful activities. |
| **Emotional Well-being** | Emotional representations (fear, anger, sadness about having Parkinson’s disease) directly affect psychological quality of life and mood stability. | Catastrophising and symptom-related fear may increase anxiety and perceived distress during symptom fluctuations. |
| **Physical Functioning** | Low perceived personal control may reduce motivation to engage in physiotherapy, exercise, or self-management behaviours. | Avoidance of movement due to fear of falling or worsening tremor can directly reduce mobility and physical independence. |
| **Fatigue-Related Quality of Life** | Belief that fatigue is uncontrollable or a sign of rapid deterioration may reduce coping efforts. | All-or-nothing behaviour (overexertion on ‘good days’ followed by exhaustion) can worsen fatigue and decrease functional capacity. |
| **Mobility and Independence** | Perceived severe consequences may increase dependency beliefs and reduce self-efficacy. | Excessive resting or activity restriction can lead to deconditioning and greater mobility limitation. |
| **Social Participation** | Beliefs about stigma or illness consequences may influence withdrawal from social roles. | Embarrassment avoidance (avoiding social settings due to visible tremor or dyskinesia) directly reduces social engagement. |
| **Cognitive Quality of Life** | Illness coherence (understanding of Parkinson’s disease) may reduce uncertainty-related stress and improve coping confidence. | Symptom focusing may amplify awareness of cognitive lapses, increasing perceived cognitive burden. |
| **Treatment Engagement** | Strong treatment control beliefs may improve medication adherence and optimism about deep brain stimulation or therapy. | Behavioural avoidance may reduce attendance at rehabilitation sessions or community activities. |
| **Fluctuation Management** | Timeline cyclical beliefs reflect awareness of motor fluctuations (‘on/off’ periods). | Behavioural responses to fluctuations (e.g., inactivity during ‘off’ periods) influence daily productivity and independence. |
| **Long-Term Adjustment** | Shapes how the person integrates Parkinson’s disease into their identity and future outlook. | Shapes daily activity patterns that accumulate over time to influence disability progression. |

#### **Supplementary material F.** Study flow diagram

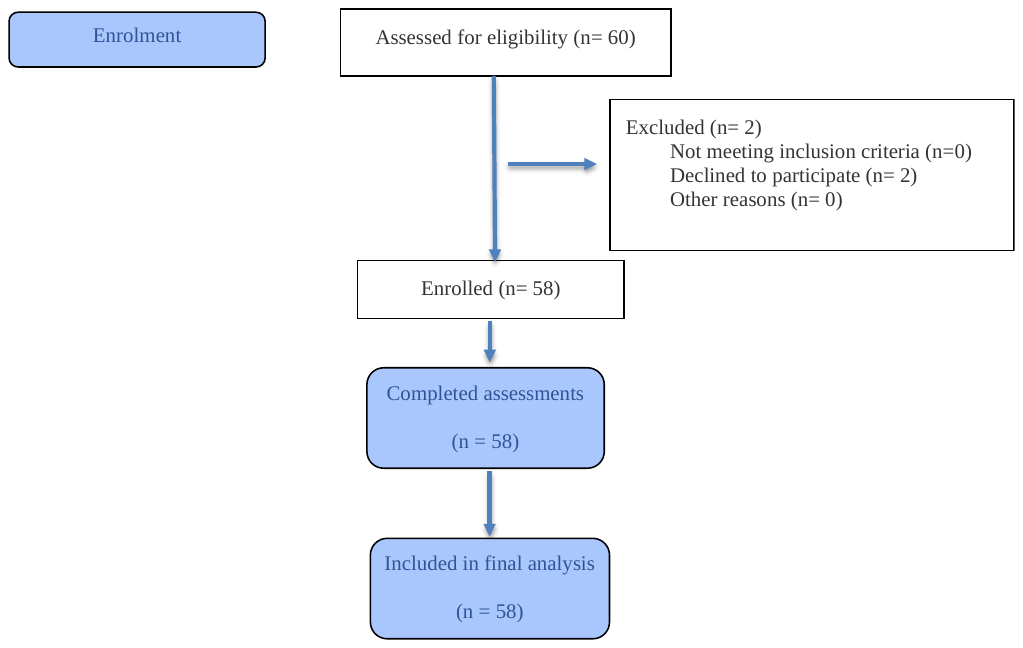

#### **Supplementary material G.** Supplementary tables (1-6) below on regression analyses

**Table 1** Hierarchical regression model comparisons predicting PDQ-39 Summary Index in full cohort

| **Model step** | **Block added** | **R^2^** | **Adj R^2^** | **ΔR^2^ vs prior** | **p (ΔF)** |
| --- | --- | --- | --- | --- | --- |
| 1 | Covariates | 0.773 | 0.735 |  |  |
| 2 | + MDS-UPDRS Part IV | 0.809 | 0.774 | 0.037 | 0.004 |
| 3a | + IPQ-R consequences | 0.834 | 0.798 | 0.024 | 0.011 |
| 3b | + CBRQ catastrophising | 0.829 | 0.792 | 0.020 | 0.025 |
| 3c | + IPQ-R consequences + CBRQ catastrophising | 0.851 | 0.815 | 0.041 | 0.004 |

R^2^, coefficient of determination; Adj R^2^, adjusted coefficient of determination; ΔR^2^, change in R^2^; p (ΔF), P value for the F-test of change in R^2^ compared with the prior model. PDQ-39, 39-item Parkinson’s Disease Questionnaire; MDS-UPDRS Part IV, Movement Disorder Society-Unified Parkinson’s Disease Rating Scale Part IV (motor complications); IPQ-R, Illness Perception Questionnaire-Revised; CBRQ, Cognitive and Behavioral Responses Questionnaire. R2, coefficient of determination; Adj R2, adjusted coefficient of determination; ΔR2, change in R2 compared with the prior model; p (ΔF), P value for the F-test of change in R2. PDQ-39, 39-item Parkinson’s Disease Questionnaire; MDS-UPDRS Part IV, Movement Disorder Society-Unified Parkinson’s Disease Rating Scale Part IV (motor complications); IPQ-R, Illness Perception Questionnaire-Revised; CBRQ, Cognitive and Behavioral Responses Questionnaire. Demographic and clinical variables, specifically, age, sex, disease duration, MDS-UPDRS Part III (motor examination), LEDD, HADS-A, HADS-D, and MoCA were entered in Block 1; motor complication severity (MDS-UPDRS Part IV) was entered in Block 2; and selected IPQ-R and CBRQ domains were entered in Block 3.

**Table 2** Collinearity diagnostics for final model in full cohort

| **Predictor** | **VIF** |
| --- | --- |
| MDS-UPDRS Part IV | 2.57 |
| Disease duration, y | 2.25 |
| HADS-D | 2.14 |
| HADS-A | 2.12 |
| CBRQ Catastrophising | 1.82 |
| Age, y | 1.71 |
| MDS-UPDRS Part III | 1.68 |
| LEDD, mg/day | 1.63 |
| IPQ-R Consequences | 1.39 |
| MoCA | 1.36 |
| Female sex | 1.35 |

VIF, variance inflation factor; MDS-UPDRS, Movement Disorder Society-Unified Parkinson’s Disease Rating Scale; Part III, motor examination; Part IV, motor complications; HADS-D, Hospital Anxiety and Depression Scale-Depression subscale; HADS-A, Hospital Anxiety and Depression Scale-Anxiety subscale; CBRQ, Cognitive and Behavioral Responses to Symptoms Questionnaire; IPQ-R, Illness Perception Questionnaire-Revised; LEDD, levodopa equivalent daily dose; MoCA, Montreal Cognitive Assessment. All VIF values <5 indicate absence of problematic multicollinearity.

**Table 3** Hierarchical regression predicting PDQ-39 Summary Index in Parkinson’s KinetiGraph subset

| **Model step** | **Block added** | **R^2^** | **Adj R^2^** | **ΔR^2^ vs prior** | **p (ΔF)** |
| --- | --- | --- | --- | --- | --- |
| 1 | Covariates | 0.810 | 0.770 |  |  |
| 2 | + MDS-UPDRS Part IV | 0.842 | 0.803 | 0.032 | 0.010 |
| 3a | + IPQ-R consequences | 0.864 | 0.826 | 0.022 | 0.020 |
| 3b | + CBRQ catastrophising | 0.854 | 0.813 | 0.012 | 0.092 |
| 3c | + IPQ-R consequences + CBRQ catastrophising | 0.873 | 0.833 | 0.031 | 0.022 |

R^2^, coefficient of determination; Adj R^2^, adjusted coefficient of determination; ΔR^2^, change in R^2^ compared with the prior model; p (ΔF), P value for the F-test of change in R^2^. PDQ-39, 39-item Parkinson’s Disease Questionnaire; MDS-UPDRS Part IV, Movement Disorder Society-Unified Parkinson’s Disease Rating Scale Part IV (motor complications); IPQ-R, Illness Perception Questionnaire-Revised; CBRQ, Cognitive and Behavioral Responses Questionnaire. Demographic and clinical variables, specifically, age, sex, disease duration, MDS-UPDRS Part III (motor examination), LEDD, HADS-A, HADS-D, and MoCA were entered in Block 1; motor complication severity (MDS-UPDRS Part IV) was entered in Block 2; and selected IPQ-R and CBRQ domains were entered in Block 3.

**Table 4** Final model coefficients in Parkinson’s KinetiGraph subset (connecting to model 3c from Supplementary Material I: Table 3)

| **Predictor** | **B** | **SE** | **β** | **95% CI** | **p** |
| --- | --- | --- | --- | --- | --- |
| Age, y | -0.269 | 0.127 | -0.172 | -0.527 to -0.012 | 0.040 |
| Female sex | -5.132 | 2.563 | -0.143 | -10.335 to 0.071 | 0.053 |
| Disease duration, y | -0.144 | 0.352 | -0.041 | -0.858 to 0.570 | 0.685 |
| MDS-UPDRS Part III | 0.117 | 0.118 | 0.084 | -0.123 to 0.356 | 0.329 |
| LEDD, mg/day | 0.022 | 0.010 | 0.193 | 0.002 to 0.042 | 0.035 |
| HADS-A | 1.409 | 0.419 | 0.314 | 0.558 to 2.261 | 0.002 |
| HADS-D | 0.652 | 0.397 | 0.153 | -0.153 to 1.457 | 0.109 |
| MoCA | -0.852 | 0.449 | -0.138 | -1.762 to 0.059 | 0.066 |
| MDS-UPDRS Part IV | 0.717 | 0.374 | 0.195 | -0.043 to 1.477 | 0.064 |
| CBRQ/CBRQ Catastrophising | 0.585 | 0.376 | 0.132 | -0.179 to 1.348 | 0.129 |
| IPQ-R Consequences | 0.785 | 0.343 | 0.170 | 0.088 to 1.482 | 0.028 |

B, unstandardized regression coefficient; SE, standard error; β, standardized regression coefficient; CI, confidence interval; LEDD, levodopa equivalent daily dose; HADS-A, Hospital Anxiety and Depression Scale-Anxiety subscale; HADS-D, Hospital Anxiety and Depression Scale-Depression subscale; MoCA, Montreal Cognitive Assessment; MDS-UPDRS, Movement Disorder Society-Unified Parkinson’s Disease Rating Scale; Part III, motor examination; Part IV, motor complications; IPQ-R, Illness Perception Questionnaire-Revised; CBRQ, Cognitive and Behavioral Responses to Symptoms Questionnaire. Model 3c corresponds to the final hierarchical model reported in Supplementary Material I Table 3, including demographic and clinical variables, MDS-UPDRS Part IV, and IPQ-R Consequences and CBRQ Catastrophising. Dependent variable: PDQ-39 Summary Index.

**Table 5** Comparison of participants with vs without PKG data

| **Variable** | **PKG (n=47)** | **No PKG (n=11)** | **p** |
| --- | --- | --- | --- |
| Age, y | 68.0 (62.0-76.0) | 68.0 (62.0-71.5) | 0.605 |
| Female sex, n (%) | 21 (44.7) | 5 (45.5) | 1.000 |
| Disease duration, y | 5.0 (2.0-8.0) | 3.0 (2.5-3.0) | 0.170 |
| LEDD, mg/day | 400.0 (300.0-500.0) | 400.0 (400.0-500.0) | 0.354 |
| MDS-UPDRS Part III | 45.0 (36.5-50.5) | 40.0 (36.5-53.5) | 0.968 |
| MDS-UPDRS Part IV | 9.0 (6.0-14.0) | 7.0 (4.5-10.5) | 0.146 |
| PDQ-39 SI | 32.0 (17.7-47.5) | 26.8 (19.2-34.8) | 0.362 |
| HADS-A | 8.0 (4.0-12.0) | 6.0 (4.5-9.0) | 0.537 |
| HADS-D | 7.0 (5.0-10.5) | 5.0 (2.0-9.0) | 0.146 |
| MoCA | 26.0 (24.0-27.0) | 26.0 (25.0-28.0) | 0.516 |

Data are presented as median (interquartile range) unless otherwise indicated. Continuous variables were compared using the Mann-Whitney U test; sex was compared using Fisher’s exact test. PKG, Parkinson’s KinetiGraph; LEDD, levodopa equivalent daily dose; MDS-UPDRS, Movement Disorder Society-Unified Parkinson’s Disease Rating Scale; Part III, motor examination; Part IV, motor complications; PDQ-39 SI, 39-item Parkinson’s Disease Questionnaire Summary Index; HADS-A, Hospital Anxiety and Depression Scale-Anxiety subscale; HADS-D, Hospital Anxiety and Depression Scale-Depression subscale; MoCA, Montreal Cognitive Assessment.

**Table 6** Parkinsons’s KinetiGraph dyskinesia metric (DK_50) as motor predictor of PDQ-39 Summary Index

| **Step** | **Block added** | **R^2^** | **ΔR^2^ vs prior** | **p (ΔF)** |
| --- | --- | --- | --- | --- |
| 1 | Covariates | 0.810 |  |  |
| 2 | + DK_50 (PKG dyskinesia) | 0.823 | 0.013 | 0.110 |
| 3 | + IPQ-R consequences + catastrophising | 0.861 | 0.038 | 0.015 |

R^2^, coefficient of determination; ΔR^2^, change in R^2^ compared with the prior model; p (ΔF), P value for the F-test of change in R^2^. PDQ-39, 39-item Parkinson’s Disease Questionnaire; PKG, Parkinson’s KinetiGraph; DK_50, median dyskinesia score derived from PKG; IPQ-R, Illness Perception Questionnaire-Revised; CBRQ, Cognitive and Behavioral Responses Questionnaire. Covariates (Block 1) included age, sex, disease duration, Movement Disorder Society-Unified Parkinson’s Disease Rating Scale Part III (motor examination), levodopa equivalent daily dose, Hospital Anxiety and Depression Scale-Anxiety and -Depression subscale scores, and Montreal Cognitive Assessment score. In Block 2, DK_50 was entered in place of Movement Disorder Society-Unified Parkinson’s Disease Rating Scale Part IV (motor complications). In Block 3, IPQ-R Consequences and CBRQ Catastrophising were entered simultaneously. The change in R^2^ for DK_50 was not statistically significant after covariate adjustment.

#### **Supplementary material H.** Adjusted (partial) correlations between clinical, psychological, and digital motor measures and health-related quality of life

| **Variable** | **Partial ρ with PDQ-39 SI (N=58)** | **P value** | **Partial ρ with PDQ-39 SI (N=47)** | **P value** |
| --- | --- | --- | --- | --- |
| **Motor complications (clinical)** |  |  |  |  |
| MDS-UPDRS Part IV | 0.356 | 0.011 | 0.405 | 0.011 |
| **Motor complications (PKG)** |  |  |  |  |
| DK_50 | - | - | 0.007 | 0.964 |
| FDS | - | - | -0.172 | 0.246 |
| % Time Immobile | - | - | 0.005 | 0.975 |
| **CBRQ subscales** |  |  |  |  |
| Fear Avoidance | 0.297 | 0.039 | 0.199 | 0.180 |
| Catastrophising | 0.362 | 0.010 | 0.277 | 0.059 |
| Damage Beliefs | 0.430 | 0.002 | 0.525 | 0.001 |
| Embarrassment Avoidance | 0.283 | 0.049 | 0.216 | 0.145 |
| Symptom Focusing | 0.214 | 0.142 | 0.204 | 0.169 |
| All-or-Nothing Behaviour | 0.352 | 0.013 | 0.377 | 0.022 |
| Avoidance/Resting Behaviour | 0.324 | 0.025 | 0.212 | 0.153 |
| **IPQ-R Part 2 domains** |  |  |  |  |
| Timeline (Chronic) | 0.096 | 0.514 | 0.100 | 0.503 |
| Consequences | 0.247 | 0.089 | 0.358 | 0.029 |
| Personal Control | -0.038 | 0.795 | 0.056 | 0.707 |
| Treatment Control | 0.278 | 0.048 | 0.226 | 0.127 |
| Illness Coherence | 0.288 | 0.042 | 0.248 | 0.092 |
| Timeline Cyclical | 0.296 | 0.039 | 0.305 | 0.065 |
| Emotional Representation | 0.246 | 0.091 | 0.073 | 0.626 |

Effect sizes are presented as partial Spearman’s rank correlation coefficients (ρ) between each variable and the PDQ-39 Summary Index (SI), adjusted for age, sex, disease duration, MDS-UPDRS Part III, levodopa equivalent daily dose (LEDD), Hospital Anxiety and Depression Scale anxiety (HADS-A) and depression (HADS-D) subscales, and Montreal Cognitive Assessment (MoCA). Partial Spearman correlations were implemented by rank-transforming variables and computing partial correlations on ranks. N = 58 for the full cohort and N = 47 for the PKG subset; N indicates complete-case data for outcome, predictor, and covariates in each analysis. The PDQ-39 SI was calculated as the mean of the eight transformed PDQ-39 domain scores (0-100), with higher scores indicating poorer health-related quality of life. Dashes indicate variables not available in the full cohort. All tests were two-sided. P values are reported for descriptive purposes only. No formal adjustment was made for multiple comparisons.

Abbreviations: PDQ-39 SI, Parkinson’s Disease Questionnaire-39 Summary Index; MDS-UPDRS, Movement Disorder Society-Unified Parkinson’s Disease Rating Scale; CBRQ, Cognitive and Behavioural Responses to Symptoms Questionnaire; IPQ-R Part 2, Illness Perception Questionnaire-Revised Part 2; PKG, Parkinson’s KinetiGraph; DK_50, median dyskinesia score; FDS, fluctuation dyskinesia score; % time immobile, percentage of time immobile.

#### **Supplementary material I.** Spearman Correlations Between Cognitive-Behavioural Responses and Illness Perceptions

**Table A** Correlations between cognitive-behavioural responses and illness perceptions in full cohort

| **Participants**  **(N = 58)** | **IPQ-R Part 2 domains** |  |  |  |  |  |  |
| --- | --- | --- | --- | --- | --- | --- | --- |
| **CBRQ Subscales** | **Timeline (Chronic)** | **Consequences** | **Personal Control** | **Treatment Control** | **Illness Coherence** | **Timeline Cyclical** | **Emotional Representation** |
| **Fear Avoidance** | 0.151 (0.256) | 0.106 (0.427) | 0.058 (0.667) | 0.358 (0.006) | 0.154 (0.247) | 0.213 (0.108) | 0.381 (0.003) |
| **Catastrophising** | 0.175 (0.190) | 0.213 (0.108) | 0.062 (0.645) | 0.298 (0.023) | 0.304 (0.020) | 0.368 (0.004) | 0.707 (<0.001) |
| **Damage Beliefs** | 0.048 (0.723) | 0.132 (0.323) | 0.054 (0.690) | 0.283 (0.031) | 0.300 (0.022) | 0.407 (0.002) | 0.358 (0.006) |
| **Embarrassment Avoidance** | -0.074 (0.580) | 0.153 (0.252) | -0.169 (0.206) | 0.155 (0.244) | 0.282 (0.032) | 0.206 (0.120) | 0.422 (0.001) |
| **Symptom Focusing** | -0.077 (0.567) | 0.372 (0.004) | 0.012 (0.931) | 0.224 (0.091) | 0.419 (0.001) | 0.305 (0.020) | 0.588 (<0.001) |
| **All-or-Nothing Behaviour** | -0.036 (0.789) | 0.138 (0.301) | -0.033 (0.807) | 0.371 (0.004) | 0.114 (0.394) | 0.379 (0.003) | 0.481 (<0.001) |
| **Avoidance/Resting Behaviour** | 0.238 (0.072) | 0.121 (0.364) | 0.152 (0.254) | 0.320 (0.014) | 0.327 (0.012) | 0.286 (0.029) | 0.460 (<0.001) |

Effect sizes are presented as Spearman’s rank correlation coefficients (ρ). N = 58 for all analyses. N indicates the number of participants with available data for each analysis. All statistical tests were two-sided. P values <0.05 are reported for descriptive purposes only. No formal adjustment was made for multiple comparisons.

Abbreviations: CBRQ, Cognitive and Behavioural Responses to Symptoms Questionnaire; IPQ-R Part 2, Illness Perception Questionnaire-Revised Part 2.

**Table B** Correlations between cognitive-behavioural responses and illness perceptions in Parkinson’s KinetiGraph Subset

| **Participants**  **(N = 47)** | **IPQ-R Part 2 domains** |  |  |  |  |  |  |
| --- | --- | --- | --- | --- | --- | --- | --- |
| **CBRQ Subscales** | **Timeline (Chronic)** | **Consequences** | **Personal Control** | **Treatment Control** | **Illness Coherence** | **Timeline Cyclical** | **Emotional Representation** |
| **Fear Avoidance** | 0.018 (0.906) | 0.013 (0.932) | 0.112 (0.453) | 0.326 (0.025) | 0.102 (0.496) | 0.183 (0.217) | 0.351 (0.016) |
| **Catastrophising** | 0.213 (0.151) | 0.166 (0.266) | 0.114 (0.444) | 0.269 (0.067) | 0.277 (0.059) | 0.339 (0.020) | 0.729 (<0.001) |
| **Damage Beliefs** | 0.079 (0.600) | 0.042 (0.779) | 0.043 (0.773) | 0.258 (0.080) | 0.242 (0.101) | 0.427 (0.003) | 0.354 (0.015) |
| **Embarrassment Avoidance** | -0.109 (0.467) | 0.103 (0.491) | -0.150 (0.314) | 0.088 (0.557) | 0.241 (0.102) | 0.138 (0.353) | 0.360 (0.013) |
| **Symptom Focusing** | -0.078 (0.603) | 0.297 (0.043) | 0.029 (0.846) | 0.193 (0.193) | 0.422 (0.003) | 0.235 (0.112) | 0.594 (<0.001) |
| **All-or-Nothing Behaviour** | -0.059 (0.695) | 0.075 (0.618) | -0.035 (0.816) | 0.328 (0.024) | 0.108 (0.471) | 0.373 (0.010) | 0.479 (0.001) |
| **Avoidance/Resting Behaviour** | 0.168 (0.258) | 0.058 (0.700) | 0.267 (0.070) | 0.310 (0.034) | 0.312 (0.033) | 0.310 (0.034) | 0.471 (0.001) |

Effect sizes are presented as Spearman’s rank correlation coefficients (ρ), with two-sided p-values shown in parentheses. N = 47 for all analyses. N indicates the number of participants with available data for each correlation. P values <0.05 are reported for descriptive purposes only. No formal adjustment was made for multiple comparisons.

Abbreviations: CBRQ, Cognitive and Behavioural Responses to Symptoms Questionnaire; IPQ-R Part 2, Illness Perception Questionnaire-Revised Part 2.

#### **Supplementary material J.** Convergence between Parkinson’s KinetiGraph metrics and MDS-UPDRS Part IV

| **PKG metric** | **Spearman’s ρ with MDS-UPDRS Part IV** | **p-value** |
| --- | --- | --- |
| DK_50 (median dyskinesia score) | 0.088 | 0.557 |
| FDS (Fluctuation Dyskinesia Score) | 0.067 | 0.656 |
| % Time Immobile | 0.020 | 0.896 |

Effect sizes are presented as Spearman’s rank correlation coefficients (ρ). N = 47 for all analyses. N indicates the number of participants with available data for each correlation. All statistical tests were two-sided. P values <0.05 are reported for descriptive purposes only. No formal adjustment was made for multiple comparisons.

Abbreviations: MDS-UPDRS, Movement Disorder Society-Unified Parkinson’s Disease Rating Scale; PKG, Parkinson’s KinetiGraph; DK_50, median dyskinesia score; FDS, fluctuation dyskinesia score; % time immobile, percentage of time immobile.
